# Development of a consensus extension of the estimands framework for cluster randomised trials (CRT-estimands): results from an international Delphi study

**DOI:** 10.1101/2025.07.03.25330799

**Authors:** Brennan C Kahan, Melanie Bahti, Dongquan Bi, Frank Bretz, Gary S Collins, Andrew Copas, Michael O Harhay, Fan Li, Catherine L Auriemma

**Affiliations:** MRC Clinical Trials Unit at UCL, London, UK; Palliative and Advanced Illness Research (PAIR) Center, Philadelphia, PA, USA; Novartis Pharma AG, Basel, Switzerland; Section for Medical Statistics, Center for Medical Statistics, Informatics, and Intelligent Systems, Medical University of Vienna, Vienna, Austria; Centre for Statistics in Medicine, Nuffield Department of Orthopaedics, Rheumatology and Musculoskeletal Sciences, University of Oxford, Oxford, UK; Department of Biostatistics, Epidemiology, and Informatics, University of Pennsylvania, Philadelphia, Pennsylvania; Department of Biostatistics, Yale School of Public Health, New Haven, CT, USA; Center for Methods in Implementation and Prevention Science, Yale School of Public Health, New Haven, CT, USA; Department of Medicine, Perelman School of Medicine, University of Pennsylvania in Philadelphia, PA, USA

**Keywords:** Cluster randomised trials, estimands, ICH E9(R1), Delphi study, consensus guidelines

## Abstract

**Background:** Estimands are increasingly used in randomised trials to clarify research objectives. The ICH E9(R1) addendum sets out five attributes necessary to describe a well-defined estimand. However, the addendum was primarily developed for individually randomised trials. There is growing recognition that estimand descriptions for cluster randomised trials, where groups of individuals are randomised, may require specification of additional considerations. We conducted a Delphi study to assess stakeholder views on additional items for inclusion in a consensus extension of the ICH E9(R1) for cluster randomised trials.

**Methods:** We invited experts in estimands and cluster randomised trials to participate in a modified Delphi process to identify critical items for describing estimands in cluster randomised trials. The research team generated an initial list of eight items and definitions. Across three Delphi rounds, panellists scored items, suggested additional items, and provided open-ended rationales for responses. The consensus threshold was set as ≥70% of respondents rating an attribute as “essential” (i.e., score of ≥7 on a 9-point Likert scale) and <15% of respondents rating the item as “not important” (i.e., a score of ≤3).

**Results:** Seventy-three (52%) invited individuals participated in Round 1. Response rates were 85% in Round 2 and 95% in Round 3. Panellists included largely statisticians (62, 85%) and clinical trialists (18, 25%). After Round 1, one additional item was added for Round 2 inclusion. After Round 3, five items met consensus criteria: how individuals and clusters are weighted, population of clusters, exposure time of clusters and individuals to the intervention, whether treatment effects are marginal or cluster-specific, and handling of cluster-level intercurrent events.

**Conclusions:** This Delphi identified expert consensus around the importance of several key items for defining estimands in cluster randomised trials. These results can inform the development of consensus guidance outlining the set of attributes to describe when defining estimands for cluster randomised trials.

## Background

Since publication of the ICH E9(R1) addendum^1^, the use of estimands in randomised trials has rapidly increased^2^. An estimand is a precise description of the treatment effect a trial sets out to quantify, and can be used to enhance clarity around the trial objectives and ensure alignment between a trial’s methods to its objectives.^1, 3–8^ The ICH E9(R1) addendum sets out five attributes that should be described in order to have a well-defined estimand (population, treatment conditions, endpoint, population-level summary measure, and strategies to handle intercurrent events).^1^

However, the ICH E9(R1) addendum was primarily developed for individually randomised trials. Cluster randomised trials involve randomising groups of individuals, such as hospitals, schools, or villages, between treatments.^9–11^ With the increased use of estimands, there is growing recognition that while the five attributes specified in the addendum will be applicable for all trials, cluster randomised trials may require additional considerations in order to have a fully defined estimand.^12–20^ For instance, a notable example highlighted in the literature is how individuals and clusters are weighted.^12, 13, 19–22^ Commonly used estimators for cluster randomised trials use different weighting schemes, and the choice of which estimator is used can implicitly lead to a different estimand being targeted and hence a different size of effect.^12, 13, 20^

We therefore convened the CRT-Estimands executive committee (BCK, MB, DB, FB, GSC, AC, MOH, FL, CA), with the aim of developing a consensus extension of the ICH E9(R1) addendum for cluster randomised trials. The objective of this extension is to provide guidance on which attributes should be described when defining estimands in cluster randomised trials. As part of developing this guidance, we have conducted a review of published cluster randomised trials which motivated the need for new guidance^23^, and a scoping review which identified potential additional items that may be included in the guidance.^24^

In this article, we present results from a three-stage Delphi study conducted to elicit stakeholder perspectives and assess consensus on potential items to be included in the final guidance. The results from this Delphi study will be used to inform a consensus meeting, in which the final guidance will be decided.

## Methods

We conducted a three-round modified Delphi consensus process^25–27^ using similar methodology to previous international consensus studies our team has led or been involved in.^28–36^ The modified Delphi process was an iterative approach that allowed stakeholders to (1) rate items for inclusion in the guidance and explain their ratings; (2) suggest additional items to include; (3) review the ratings and explanations of other participants; and (4) subsequently revise responses based upon information learned in a prior round. This study was approved as exempt by the University of Pennsylvania IRB (protocol #856778). The protocol was posted prospectively on the Open Science Framework on 16 September 2024 (https://osf.io/a5kdy/). This study is reported following the ACCORD guidelines for consensus-based research (Supplement).^37^

## Participant recruitment

We convened a panel of stakeholders with expertise in cluster randomised trials and/or estimands, including statisticians, methodologists, clinicians, and other healthcare professionals, and researchers from related fields.

Potential experts were identified through the authorship team’s professional networks, review of author lists from a recent scoping review^24^, and through a process of “snowballing” in which invited experts could recommend additional colleagues for participation in the study.^38^ Members of the executive committee were invited to participate in the Delphi. Prospective panellists were invited to participate directly by the authors via email and were informed that completing the study surveys indicated informed consent to participate.

## Survey design and administration

The overarching objective of this Delphi study was to identify items with expert-consensus for consideration of inclusion in a consensus extension of the ICH E9(R1) addendum for cluster randomised trials. An initial list of items was identified through a scoping review of literature addressing estimands in cluster randomised trials.^24^ Descriptions and explanations of each item were developed by the executive committee iteratively through internal piloting for readability of explanations (**Table 1**).

**Table 1.**
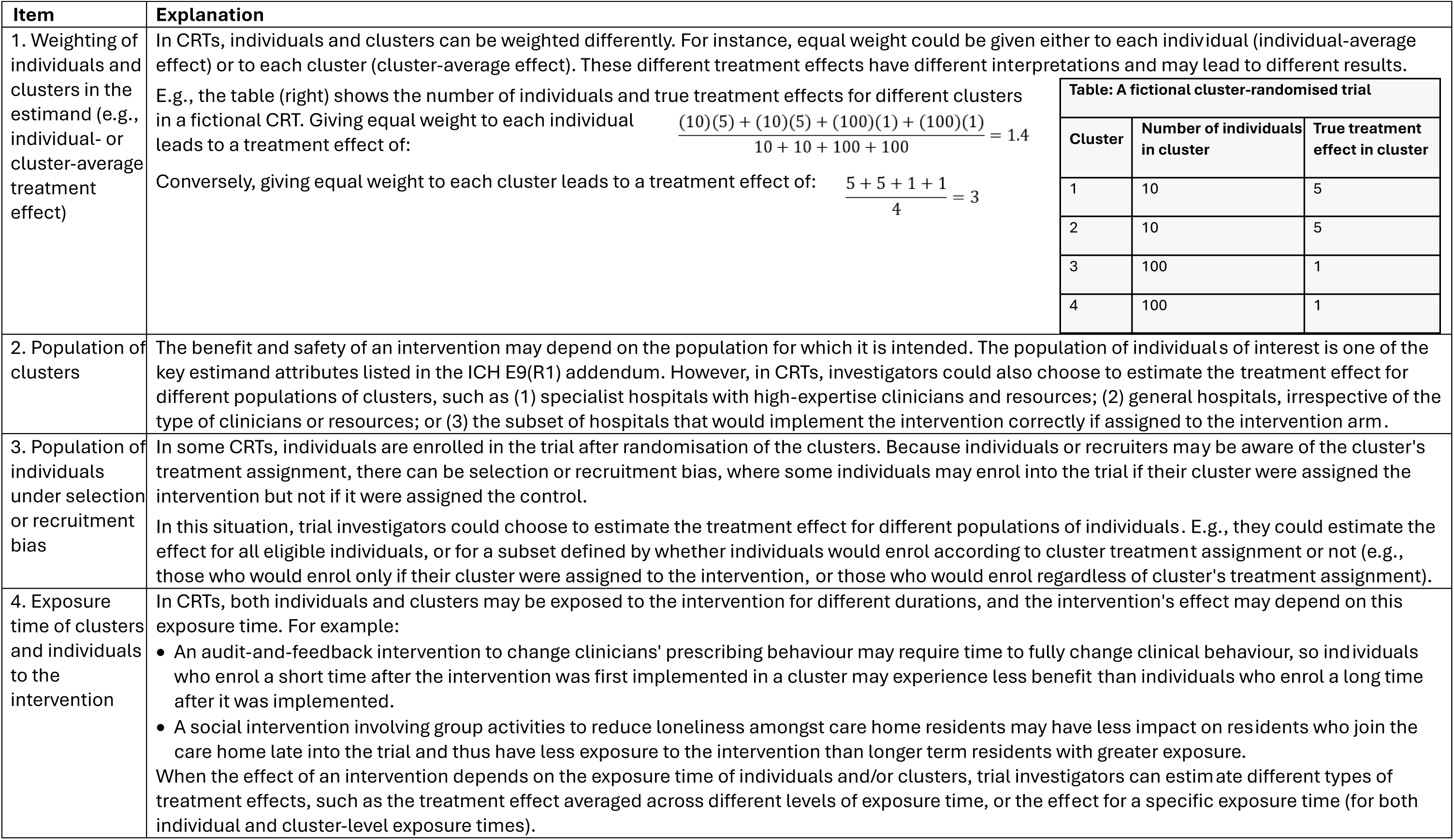
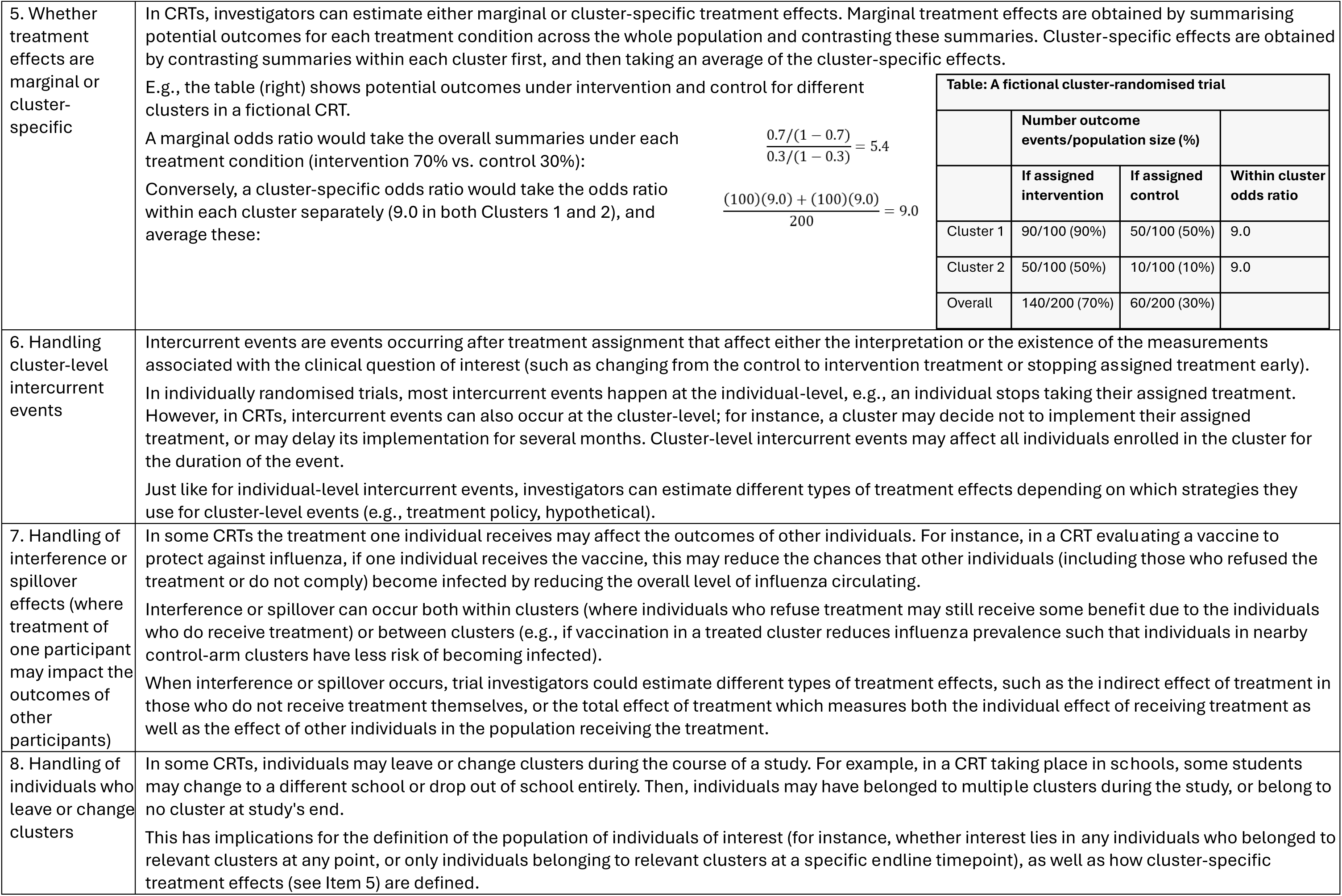
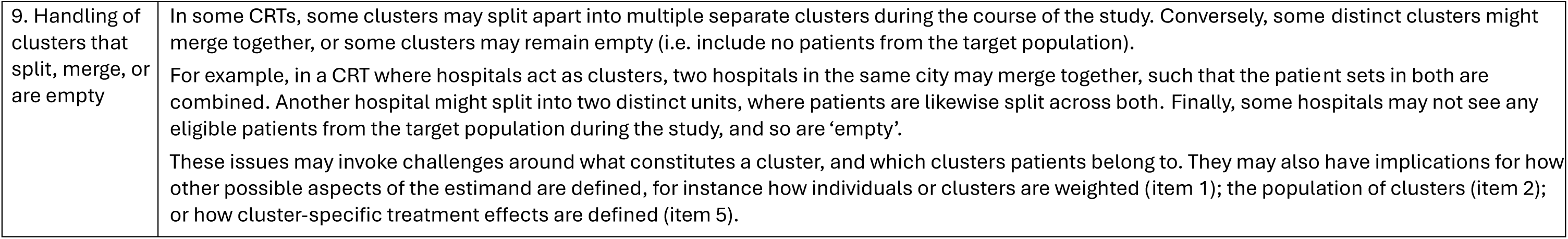
All Items and Definitions

Anonymity of individual panellists’ responses was maintained throughout the Delphi process. The REDCap online survey platform hosted at the University of Pennsylvania was used to collect demographic information as well as to administer the Delphi surveys.^39, 40^ Members of the executive group piloted the surveys to ensure functionality and clarity of questions. Participants received up to three reminder emails to complete each survey round. The Delphi study was conducted between 16 October 2024, and 10 February 2025.

## Round 1

In Round 1, participants reviewed the initial list of eight items and the definitions prepared by the executive committee. Participants were asked to rate the importance of each item and, optionally, explain their ratings for each in open-ended text. After rating the importance of the initial eight items, participants were invited to suggest additional items to be considered for inclusion in the guidance. Round 1 was conducted between 16 and 31 October 2024.

## Round 2

Following the completion of Round 1, participant responses were compiled and summarised in an executive summary distributed to panellists by email along with an invitation to complete Round 2 (Supplement). Summary ratings were presented graphically. A selection of comments from participants’ free-text explanations were summarised by the executive committee to ensure that comments across a range of ratings were represented and to communicate any substantive concerns or reasoning raised by participants. A full list of all comments was available to participants by hyperlink. Participants were asked to review the summary prior to initiating the Round 2 survey.

The Round 2 survey included the initial eight items that were rated in Round 1 and an additional ninth item that was developed based on suggestions made by multiple participants in Round 1. Round 2 included an importance-rating question for all nine items. For the first eight items, participants were able to view their own rating and the average rating from Round 1, a graphical representation of Round 1 ratings, and the selected comments from the executive summary embedded within the survey platform (in addition to their inclusion in the executive summary). For the ninth item, which was being rated for the first time, participants could optionally provide an explanation of their importance rating in open-ended text. Only individuals who completed Round 1 were invited to complete Round 2. Round 2 was conducted between 20 November and 9 December 2024.

## Round 3

Following Round 2, participant responses were again compiled and summarised in an executive summary distributed to panellists by email along with an invitation to complete Round 3 (**Supplement**). The summary again included a graphical representation of ratings for the item and a representative selection of participant comments from Round 2. A full list of comments was available to participants by hyperlink. Participants were asked to review the summary prior to completing the Round 3 survey.

The Round 3 survey included only the new item from Round 2 so it could be rated a second time. Participants were able to view their own rating and the average rating from Round 2, a graphical representation of Round 2 ratings, and the selected comments from the executive summary embedded within the survey platform (in addition to their inclusion in the executive summary). Participants were asked to rate the importance of item 9 and invited to provide any additional comments about the consensus extension of the ICH E9(R1) addendum for cluster randomised trials in open-ended text. Only individuals who completed Round 2 were invited to complete Round 3. Round 3 was conducted between 24 January and 10 February 2025.

## Statistical reporting and analysis

Response rates were defined as the proportion of invited panellists who completed each survey. Quantitative responses were summarised using descriptive statistics. The importance of an item was rated on a 9-point Likert scale ranging from 1 (“Not Important”) to 9 (“Critical”). Participants could also select “Unable to rate”. The consensus threshold for considering an item as essential was set a priori as ≥70% of participants rating an item as “Critical” (i.e., rating ≥7) and <15% of respondents rating the item as “Not Important” (i.e., rating of ≤3). Consensus threshold for considering an item as not essential for inclusion in the guidance was set a priori as ≥70% of participants rating an item as “Not Important” (i.e., rating ≤3) and <15% rating it as “Critical” (i.e., rating ≥7). Any other combination of ratings indicated no consensus about whether or not to include the item. Similar consensus definitions have been used in prior studies to ensure that an item will not achieve consensus if a subset of stakeholders commonly rates it as not important.^28, 29, 41, 42^ Final assessments of consensus were determined after an item had been rated twice (after Round 2 for Items 1-8 and after Round 3 for Item 9). Free-text responses were analysed using comparison techniques across the range of observed ratings within an item and across items by concerns or reasoning raised by participants.

## Results

Invitations to participate in the Delphi were initially sent to 114 individuals (**Figure 1**). From snowball sampling, 21 participants made a total of 50 suggestions for additional people to invite to the Delphi. Of the 50 suggestions, there were 42 unique individuals recommended, 13 of whom (31%) had already been included in the initial round of invitations. Twenty-seven of the suggested 29 additional unique individuals were subsequently invited to participate (two of the unique suggestions were made too close to the survey deadline for invitations to be sent). A total of 141 individuals received invitations to complete Round 1 of the Delphi.

**Figure 1.**
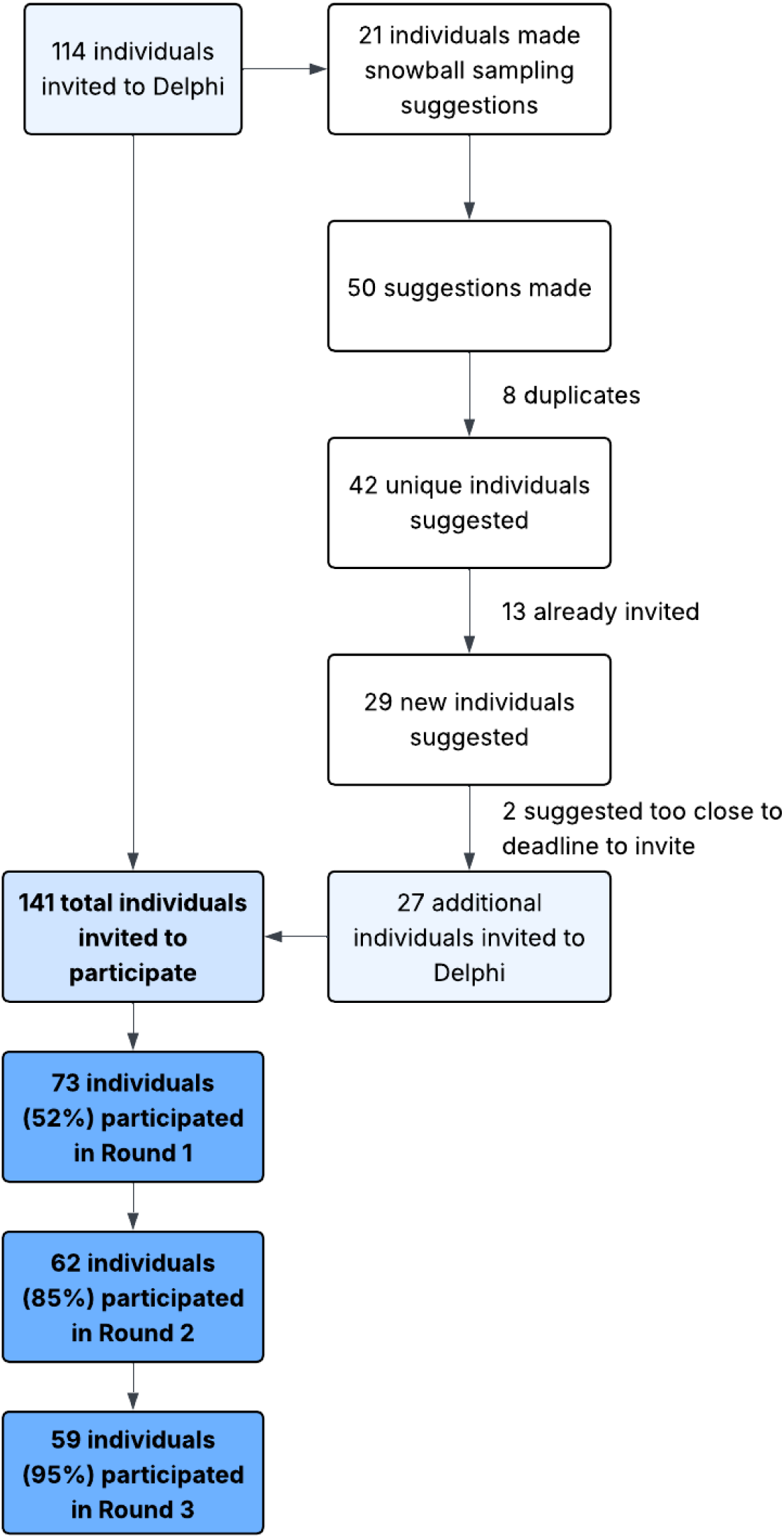
**Participant recruitment and retention**. Only individuals who completed Round 1 were invited to participate in Round 2 and only individuals who completed Round 2 were invited to participate in Round 3.

Seventy-three individuals participated in Round 1 of the Delphi (response rate 52%) and are therefore considered members of the panel **(Table 2)**. Most participants identified as a statistician (62, 85%) and/or a clinical trialist (18, 25%) and had prior experience in cluster randomised trials (60, 82%). Half or more of participants reported at least 10 years of experience in clinical trials (37, 50%) and had been involved in six or more clinical trials (37, 51%). Participants reported residing in North America (33, 45%), Europe (26, 36%), and Australia/Oceania (11, 15%).

**Table 2.**
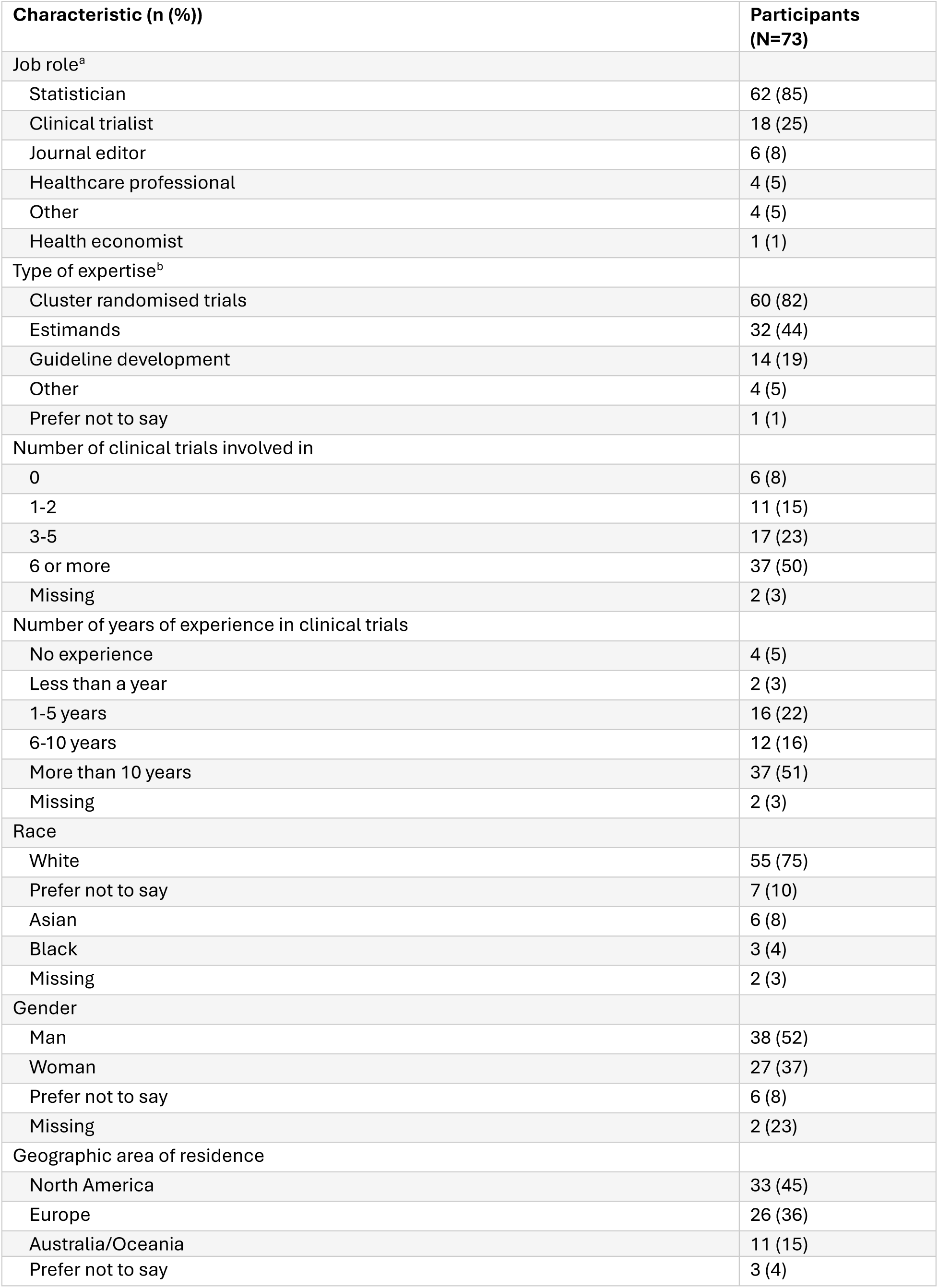
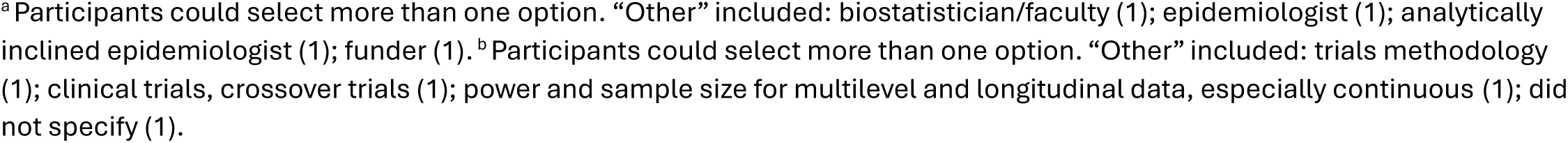
Participant Characteristics.

## Round 1

In Round 1 of the Delphi, participants rated eight items for potential inclusion in an extension of the ICH E9(R1) addendum for cluster randomised trials. The highest-rated items were “Weighting of individuals and clusters in the estimand” (mean rating 8.22, standard deviation (SD) 1.36); “Whether treatment effects are marginal or cluster-specific” (7.73, 1.55); and “Handling of cluster-level intercurrent events” (7.51, 1.69).

The lowest-rated items were “Handling of interference or spillover effects” (6.46, 2.18); “Exposure time of clusters and individuals to the intervention” (6.77, 2.12); and “Handling of individuals who leave or change clusters” (6.80, 1.73) **(Table 3)**. Review of open-ended responses suggested that Delphi participants felt these items were relevant to only a minority of cluster randomised trials, that they were already covered within the existing five ICH E9(R1) attributes, or that they were not directly related to estimands (**Table S1).**

**Table 3.** Importance ratings across rounds.

| Item | Round 1 |  |  | Round 2 |  |  | Round 3 |  |  |
| --- | --- | --- | --- | --- | --- | --- | --- | --- | --- |
|  | n* | Mean | SD | n* | Mean | SD | n* | Mean | SD |
| 1. Weighting of individuals and clusters in the estimand | 73 | 8.22 | 1.36 | 62 | 8.53 | 0.78 | n/a |  |  |
| 2. Population of clusters | 71 | 7.25 | 1.95 | 62 | 7.56 | 1.54 | n/a |  |  |
| 3. Population of individuals under selection or recruitment bias | 71 | 7.07 | 1.76 | 62 | 6.66 | 1.66 | n/a |  |  |
| 4. Exposure time of clusters and individuals to the intervention | 73 | 6.77 | 2.12 | 62 | 7.21 | 1.59 | n/a |  |  |
| 5. Whether treatment effects are marginal or cluster-specific | 70 | 7.73 | 1.55 | 62 | 7.95 | 1.32 | n/a |  |  |
| 6. Handling of cluster-level intercurrent events | 72 | 7.51 | 1.69 | 62 | 7.97 | 1.21 | n/a |  |  |
| 7. Handling of interference or spillover effects | 70 | 6.46 | 2.18 | 61 | 6.28 | 1.76 | n/a |  |  |
| 8. Handling of individuals who leave or change clusters | 70 | 6.80 | 1.73 | 62 | 6.84 | 1.52 | n/a |  |  |
| 9. Handling of clusters that split, merge, or are empty | n/a |  |  | 61 | 5.84 | 1.89 | 59 | 5.19 | 1.41 |
| *n = number of ratings provided in a given round (excludes missing data due to non-response and respondents who selected “unable to rate”). |  |  |  |  |  |  |  |  |  |
| n/a, not assessed; SD, standard deviation |  |  |  |  |  |  |  |  |  |

Three items met the threshold for consensus as essential for inclusion after Round 1: “Weighting of individuals and clusters in the estimand”; “Whether treatment effects are marginal or cluster-specific”; and “Handling of cluster-level intercurrent events” (**Table 4**).

**Table 4.**
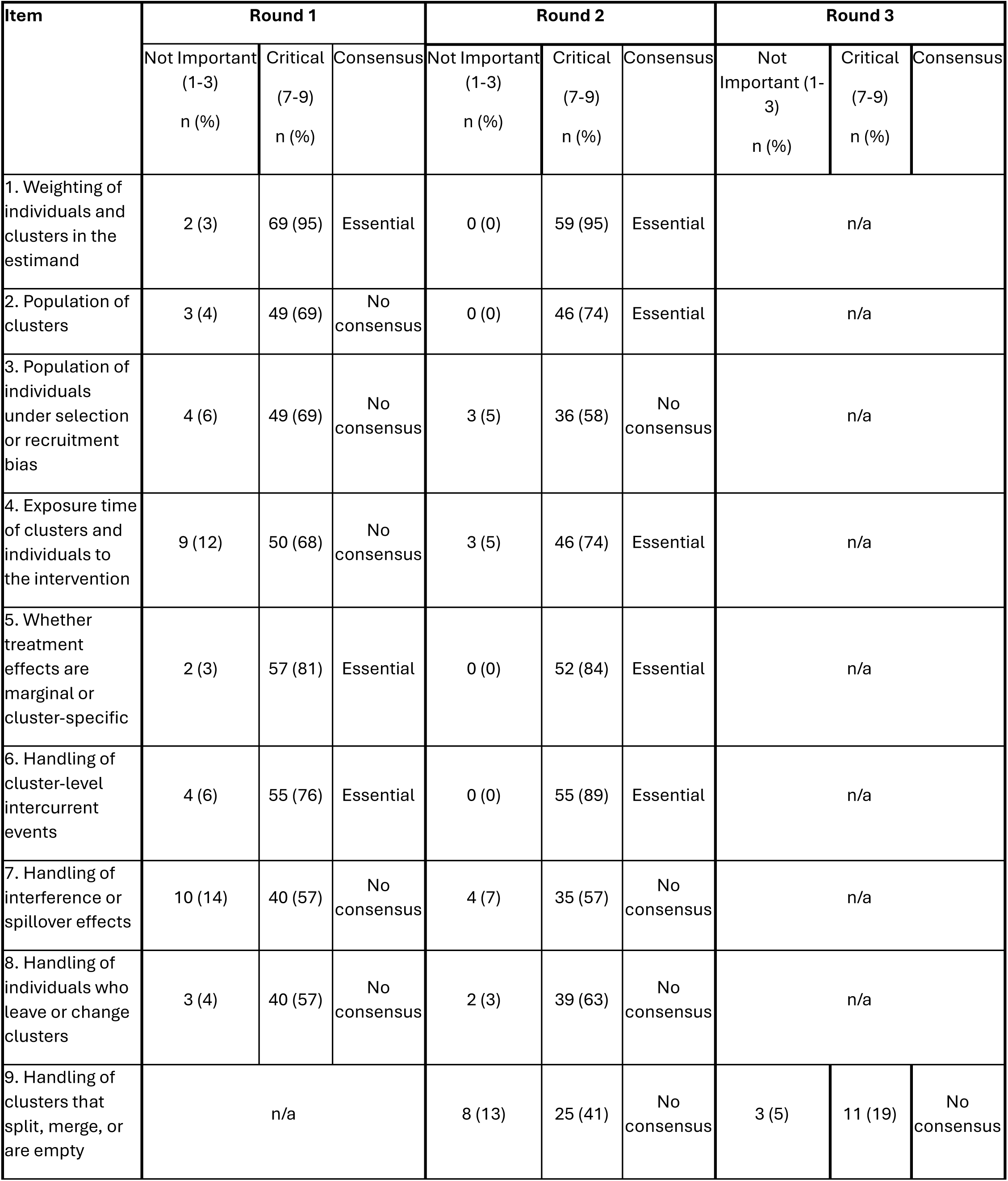
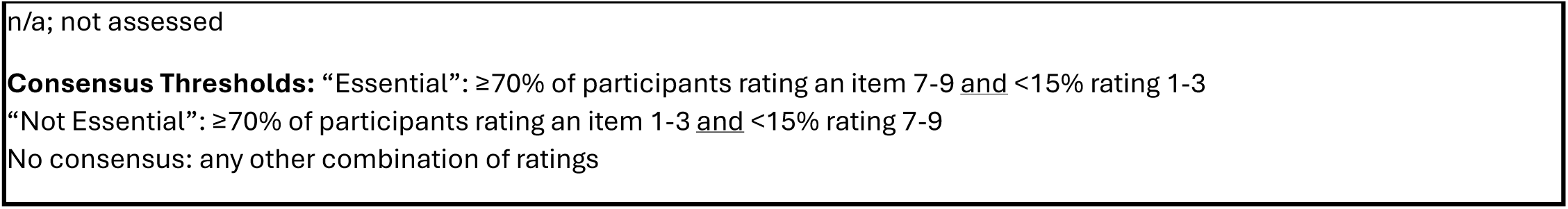
Consensus assessments across rounds.

Participants’ open-ended comments were reviewed for suggestions of topics not already included in the initial list of items. From these suggestions, a single new item was proposed: “Handling of clusters that split, merge, or are empty” (**Table 1**).

## Round 2

Round 2 was completed by 62 individuals (85% response rate). In Round 2, participants rated all items from Round 1 as well as one newly added item, “Handling of clusters that split, merge, or are empty”. Five items officially met consensus as essential after Round 2: “How individuals and clusters are weighted in the estimand”; “Population of clusters”; “Exposure time of clusters and individuals to the intervention”; “Whether treatment effects are marginal or cluster-specific”; and “Strategies for handling cluster-level intercurrent events” **(Table 4)**.

The new item rated for the first time in Round 2, “Handling of clusters that split, merge, or are empty”, received a mean rating of 5.84 (SD 1.89), with 25 (41%) rating it as “Critical” and 8 (13%) rating as “Not Important”. In open-ended comments, multiple participants noted that this was an uncommon occurrence (**Table S1**).

## Round 3

Round 3 was completed by 59 individuals (95% response rate). The only item rated in this round was “Handling of clusters that split, merge, or are empty”. Eleven (19%) participants rated this item as “Critical” and three (5%) rated it as “Not Important”; there was no consensus as to whether it was essential or not essential to include in the guidance **(Table 3)**.

## Discussion

Though precise definitions of estimands are increasingly adopted in randomised trials, concerns have been raised that the five attributes outlined in the ICH E9(R1) addendum may not be sufficient for a clear and comprehensive estimand definition in cluster randomised trials. Based on recent work evaluating the use of estimands in cluster randomised trials, there is a clear need for updated guidance in this area.^23^ To fill this gap, we recently convened a group to develop a consensus extension of the ICH E9(R1) addendum for cluster randomised trials.

In a previous scoping review, we identified eight potential additional items that could be used to help define the estimand for cluster randomised trials.^24^ In the Delphi survey described in this article, we asked expert stakeholders to rate the importance of each of these proposed items, as well as provide feedback to explain their rating, or suggest additional items for consideration in the guidance.

Because one additional item was added during the Delphi, based on participant suggestions, stakeholders rated nine items in total. Of these, five items achieved consensus at the end of the Delphi (how individuals and clusters are weighted; population of clusters; exposure time of individuals and clusters; whether effects are marginal or cluster-specific; and strategies for handling cluster-level intercurrent events). Key themes that emerged from participant feedback on these items were that they were viewed both as applicable to a large percentage of cluster randomised trials and were essential for proper interpretation of trial results.

The remaining four items did not achieve consensus, either for their inclusion or their exclusion. Common reasons given by Delphi participants for low scores were that items were only relevant to a small percentage of cluster randomised trials; that items were already covered by the existing ICH E9(R1) attributes; or that items were not relevant to the estimand and would be better handled elsewhere (e.g., as part of the description of planned statistical methods).

These results may indicate a preference from stakeholders for briefer guidance which contains only the most essential items, rather than a more inclusive approach which expands the number of items included in the guidance. The results from this Delphi will be used to inform discussions at this project’s consensus meeting, in which the final guidance will be decided. Notably, all items assessed in the Delphi will be carried forward to the consensus meeting, as no items reached consensus as not important.

A notable strength of this study was the high response rate of the Delphi. Seventy-three of 141 invited individuals participated in Round 1. Moreover, retention in subsequent rounds exceeded 80%, indicating a high degree of interest in this project. Furthermore, Delphi participants were on average highly experienced (e.g., the majority had >10 years’ experience in clinical trials and had been involved in >5 trials), indicating that the Delphi results can be seen to represent the views of experts in this area. An additional strength is our inclusion of snowball sampling as a method to expand representativeness of the experts invited to participate. Notably, 30% of those individuals recommended by participants were already included in our invitation lists, suggesting substantial coverage of the experts in this field.

A limitation of this work is that, like with all Delphi surveys, the results may not be generalisable beyond those individuals who participated. Furthermore, a number of the items in the Delphi involve complex or technical concepts – although we are confident in the expertise of the participants included in this Delphi, we did not formally assess for comprehension of each proposed item. As such, we cannot rule out that some participants may have understood items differently from one another.

In conclusion, the results from this Delphi provide strong evidence around the importance of multiple key items when defining estimands for cluster randomised trials. These results will be used in the development of consensus guidance outlining the set of attributes that should be described when defining estimands for cluster randomised trials.

## Data Availability

The datasets used and/or analysed during the current study are available from the senior author upon reasonable request.

## Ethics approval and consent to participate

This study was approved as exempt by the University of Pennsylvania IRB (protocol #856778).

## Authors’ contributions

All authors conceived of the study. All authors participated in the design of the study. BCK, MB, and CA conducted data acquisition. All authors analysed and interpreted the data, revised the manuscript critically for important intellectual content, approved the final manuscript, and agreed to be accountable for its overall content.

### Acknowledgements

We thank the Delphi panellists who participated in this study for their time and perspectives: Agnès Caille; Rebecca Andridge; Laura B. Balzer; Andrew W Brown; Ashley Buchanan; Mike Campbell; Siobhan Creanor; Catherine M. Crespi; Anurika De Silva; Andrew Forbes; Bruno Giraudeau; Deborah H. Glueck; Richard J Hayes; Jennifer Hellier; Karla Hemming; Joanna Hindley; James P. Hughes; Lee Kennedy-Shaffer; Kenneth M. Lee; Clémence Leyrat; David P. MacKinnon; Joanne McKenzie; Lynne Moore; Tim P. Morris; Keith E. Muller; David M. Murray; Stephen Nash; Kate A. Nelson; Joshua R. Nugent; Sherri L. Pals; Pierre Poupin; Joseph S. Ross; Anca Chis Ster; Lehana Thabane; Elizabeth L. Turner; Obioha Ukoumunne; Rebecca Walwyn; Bingkai Wang; Samuel I. Watson; Lisa Yelland

