## Supplement for "Development of a consensus extension of the estimands framework for cluster randomised trials (CRT-estimands): results from an international Delphi study"

**Online Supplement**

1. ACCORD Guidelines Checklist for reporting consensus methods
2. Round 1 Executive Summary
3. Round 2 Executive Summary
4. Table S1. Selected comments from executive summaries

**Accord Reporting Checklist.**

| Item No. | Section | Checklist Item (*help text*) | Page No. |
|---|---|---|---|
| T1 | **Title** | Identify the article as reporting a consensus exercise and state the consensus methods used in the title. | 1 |
| I1 | **Introduction** | Explain why a consensus exercise was chosen over other approaches. | 4 |
| I2 | | State the aim of the consensus exercise, including its intended audience and geographical scope (national, regional, global). | 4 |
| I3 | | If the consensus exercise is an update of an existing document, state why an update is needed, and provide the citation for the original document. | 4 |
| M1 | **Methods** Registration | If the study or study protocol was prospectively registered, state the registration platform and provide a link. If the exercise was not registered, this should be stated. | 5 |
| M2 | Selection of SC and/or panellists | Describe the role(s) and areas of expertise or experience of those directing the consensus exercise. | 4-5 |
| M3 | | Explain the criteria for panellist inclusion and the rationale for panellist numbers. State who was responsible for panellist selection. | 5 |
| M4 | | Describe the recruitment process (how panellists were invited to participate). | 5 |
| M5 | | Describe the role of any members of the public, patients or carers in the different steps of the study. | N/A |
| M6 | Preparatory research | Describe how information was obtained prior to generating items or other materials used during the consensus exercise. | 5 |
| M7 | | Describe any systematic literature search in detail, including the search strategy and dates of search or the citation if published already. | 5 |
| M8 | | Describe how any existing scientific evidence was summarised and if this evidence was provided to the panellists. | 5 |
| M9 | Assessing consensus | Describe the methods used and steps taken to gather panellist input and reach consensus (for example, Delphi, RAND-UCLA, nominal group technique). | 5-7 |
| M10 | | Describe how each question or statement was presented and the response options. State whether panellists were able to or required to explain their responses, and whether they could propose new items. | 6-7 |
| M11 | | State the objective of each consensus step. | 6-7 |
| M12 | | State the definition of consensus (for example, number, percentage, or categorical rating, such as 'agree' or 'strongly agree') and explain the rationale for that definition. | 7 |
| M13 | | State whether items that met the prespecified definition of consensus were included in any subsequent voting rounds. | 7 |
| M14 | | For each step, describe how responses were collected, and whether responses were collected in a group setting or individually. | 5 |
| M15 | | Describe how responses were processed and/or synthesised. | 7-8 |
| M16 | | Describe any piloting of the study materials and/or survey instruments. | 5-6 |
| M17 | | If applicable, describe how feedback was provided to panellists at the end of each consensus step or meeting. | 6-7 |
| M18 | | State whether anonymity was planned in the study design. Explain where and to whom it was applied and what methods were used to guarantee anonymity. | 5 |
| M19 | | State if the steering committee was involved in the decisions made by the consensus panel. | 5 |
| M20 | Participation | Describe any incentives used to encourage responses or participation in the consensus process. | 6 |
| M21 | | Describe any adaptations to make the surveys/meetings more accessible. | 6-7 |
| R1 | Results | State when the consensus exercise was conducted. List the date of initiation and the time taken to complete each consensus step, analysis, and any extensions or delays in the analysis. | 6-7 |
| R2 | | Explain any deviations from the study protocol, and why these were necessary. | N/A |
| R3 | | For each step, report quantitative (number of panellists, response rate) and qualitative (relevant socio-demographics) data to describe the participating panellists. | 8-10 |
| R4 | | Report the final outcome of the consensus process as qualitative (for example, aggregated themes from comments) and/or quantitative (for example, summary statistics, score means, medians and/or ranges) data. | 10-11 |

| | | | |
|---|---|---|---|
| R5 | | List any items or topics that were modified or removed during the consensus process. Include why and when in the process they were modified or removed. | N/A |
| D1 | Discussion | Discuss the methodological strengths and limitations of the consensus exercise. | 11 |
| D2 | | Discuss whether the recommendations are consistent with any pre-existing literature and, if not, propose reasons why this process may have arrived at alternative conclusions. | 11 |
| O1 | Other information | List any endorsing organisations involved and their role. | N/A |
| O2 | | State any potential conflicts of interests, including among those directing the consensus study and panellists. Describe how conflicts of interest were managed. | 14 |
| O3 | | State any funding received and the role of the funder. | 14 |

From: PLoS Med 21(1): e1004326. https://doi.org/10.1371/journal.pmed.1004326 For more information see: https://www.ismpp.org/accord

**CRT Estimands Extension Delphi: Round 1 Executive Summary**

### *Overview of Executive Summary*

This executive summary presents the results from Round 1 of the CRT Estimands Extension Delphi, with data collected from October 17-31, 2024.

**Seventy-three individuals completed or partially completed the round 1 survey (response rate of 52%).**

The summary includes:

The summary for each item includes a selection of participant comments from Round 1. For those who are interested, a document including all participant comments for all items can be accessed here.

Item 1

**How individuals and clusters are weighted in the estimand (i.e., individual- or cluster-average treatment effect).**

*Explanation:* In CRTs, individuals and clusters can be weighted differently. For instance, equal weight could be given either to each individual (individual-average effect) or to each cluster (cluster-average effect). These different treatment effects have different interpretations and may lead to different results.

### Results

73 participants responded to this item; all provided a rating.

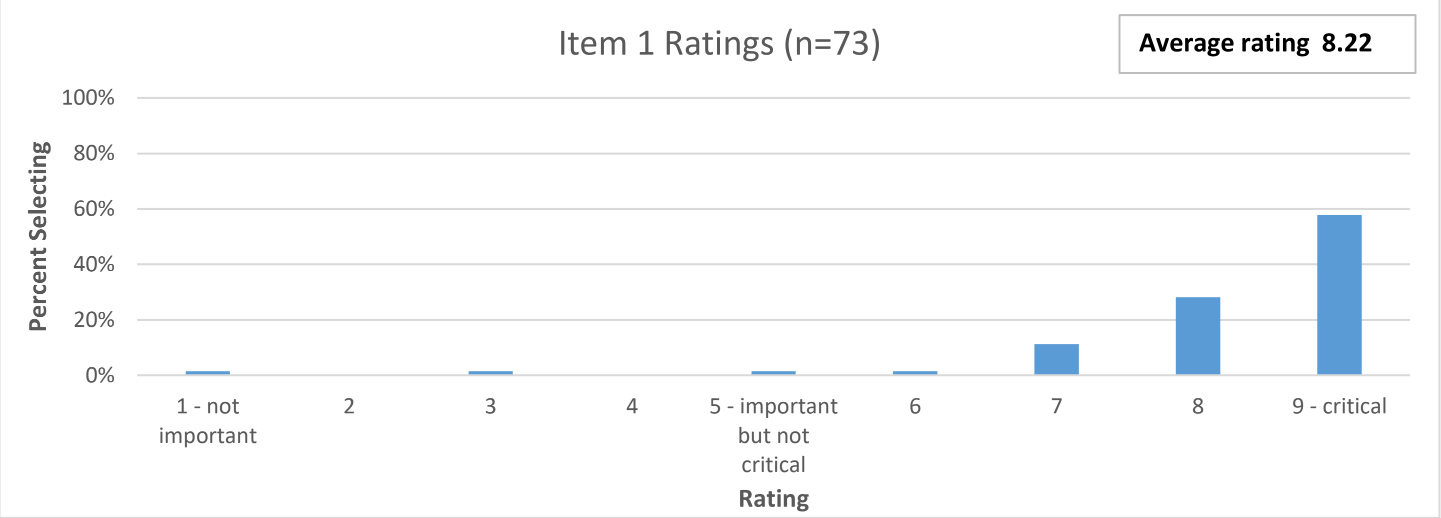

**Comments in favour of importance**

- "Absolutely critical. If clusters are of equal size, this does not matter, but this latter situation is an exception in CRTs not the rule."
- "This is fundamentally important to understanding what is being estimated, except in rare situations where the sample size is the same in all clusters."
- "It is critical because you can get quite different results depending on how individuals and clusters are weighted."
- "Critical to define as methods grow more complex; the estimand is less often implied by the analysis method than it used to be."
- "This is absolutely essential … because common analysis methods target different estimands it's essential to know which investigators want to be targeting in order to understand whether the analysis is appropriate, and what assumptions it is making."

**Comments against importance**

- Weights in analysis model more important; specifying estimand weights may indicate common analysis models not appropriate
- Only relevant if informative cluster size is present (and will not matter if cluster sizes are all equal)
- It does not seem to be something clinicians care about or are able to specify in every trial

*Item 2*

**Population of clusters**

*Explanation:* The benefit and safety of an intervention may depend on the population for which it is intended. The population of individuals of interest is one of the key estimand attributes listed in the ICH E9(R1) addendum. However, in CRTs, investigators could also choose to estimate the treatment effect for different populations of clusters, such as:

- Specialist hospitals with high-expertise clinicians and resources
- General hospitals, irrespective of the type of clinicians or resources
- The subset of hospitals that would implement the intervention correctly if assigned to the intervention arm

### Results

73 participants responded to this question; 2 selected ‘unable to rate’.

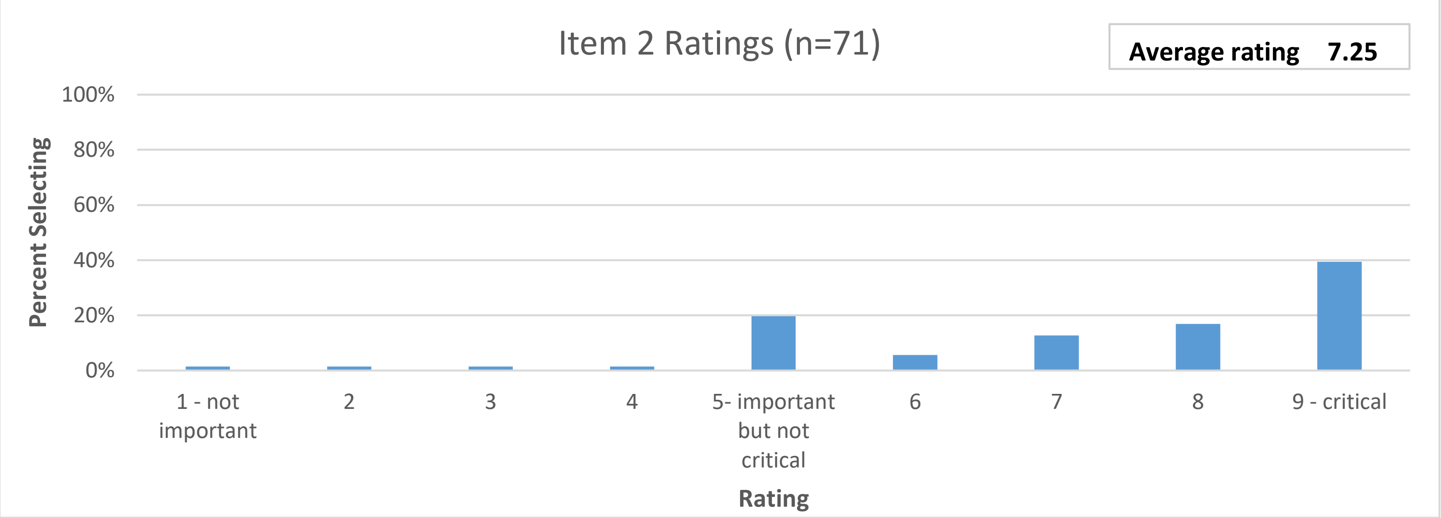

**Comments in favour of importance**

- “Both description of individuals of interest and of clusters of interest are key estimand attributes for a CRT”
- “This is important to interpreting results”
- “This is important because it affects generalisability”

**Comments against importance**

- “It seems to me that this information would already be captured in explaining the population per the existing ICH E9(R1) guidance”
- “Any trial report should describe the cluster population.”

*Item 3*
**Population of individuals under selection or recruitment bias**
*Explanation:* In some CRTs, individuals are enrolled in the trial after randomisation of the clusters. Because individuals or recruiters may be aware of the cluster's treatment assignment, there can lead to selection or recruitment bias, where some individuals may enrol into the trial if their cluster were assigned the intervention but not if it were assigned the control.

In this situation, trial investigators could choose to estimate the treatment effect for different populations of individuals. For instance, they could estimate the effect for all eligible individuals, or for a certain subset defined by whether individuals would enrol according to the cluster treatment assignment or not (e.g., those who would enrol only if their cluster were assigned to the intervention, or those who would enrol regardless of their cluster's treatment assignment).

### Results

73 participants responded to this question; 2 selected ‘unable to rate’.

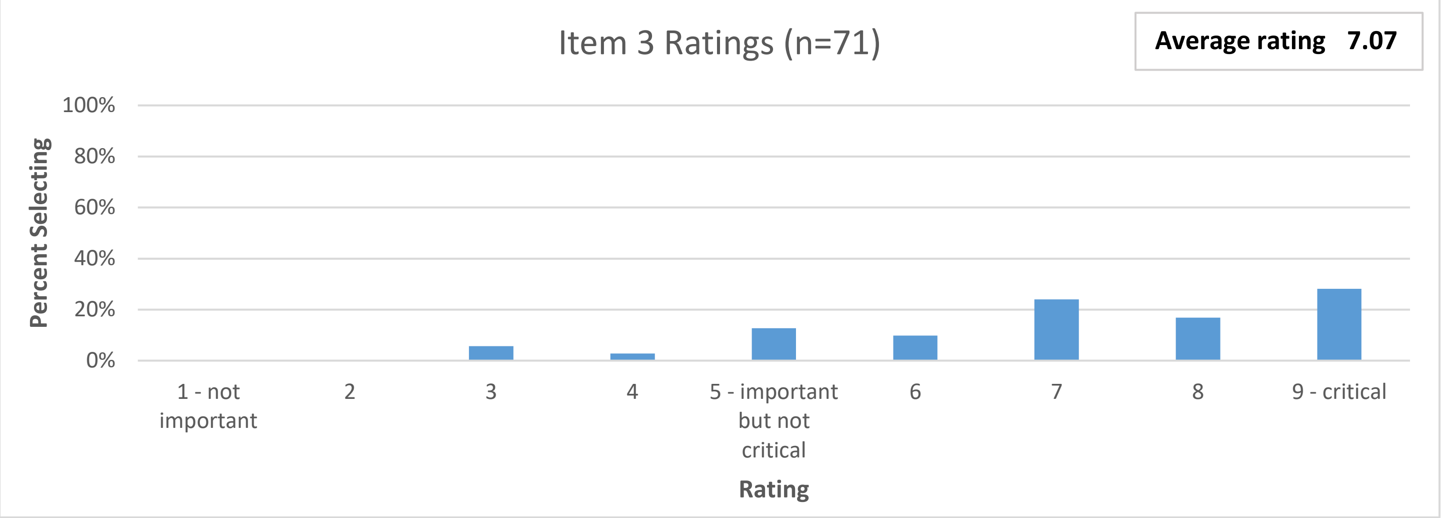

**Comments in favour of importance**

- “Important because it affects the interpretability of results”
- “This is central to understanding the ultimate effect estimate reported in the study. Without this context, making sense of results and thinking about using results in new settings is constrained.”
- “The risk for such selection or recruitment bias can be quite high and that this point should be considered very seriously in CRTs.”

**Comments against importance**

- “An important issue for the conduct of some CRTs (not all) but not necessarily an issue to do with estimands. It seems that the "clinical question of interest" would almost always relate to "all eligible individuals" since in future clinical implementation the issue of post-randomisation enrolment would not apply.”
- “While this is important for trials affected by selection/recruitment bias, it is not relevant for all trials. It is also (in principle) covered by the "Patient population" attribute. Finally, it can be a technically challenging concept, and investigators may find it challenging to implement routinely (which then makes the guidance less useful overall, if people find it difficult to use).”

*Item 4*

**Exposure time of clusters and individuals to the intervention**

*Explanation:* In CRTs, both individuals and clusters may be exposed to the intervention for different durations, and the intervention's effect may depend on this exposure time. For example:

- An audit-and-feedback intervention to change clinicians' prescribing behaviour may require time to fully change clinical behaviour, so individuals who enrol a short time after the intervention was first implemented in a cluster may experience less benefit than individuals who enrol a long time after it was implemented.
- A social intervention involving group activities to reduce loneliness amongst care home residents may have less impact on residents who join the care home late into the trial and thus have less exposure to the intervention than longer term residents with greater exposure.

When the effect of an intervention depends on the exposure time of individuals and/or clusters, trial investigators can estimate different types of treatment effects, such as the treatment effect averaged across different levels of exposure time, or the effect for a specific exposure time (for both individual and cluster-level exposure times).

### Results

73 participants responded to this question; all provided a rating.

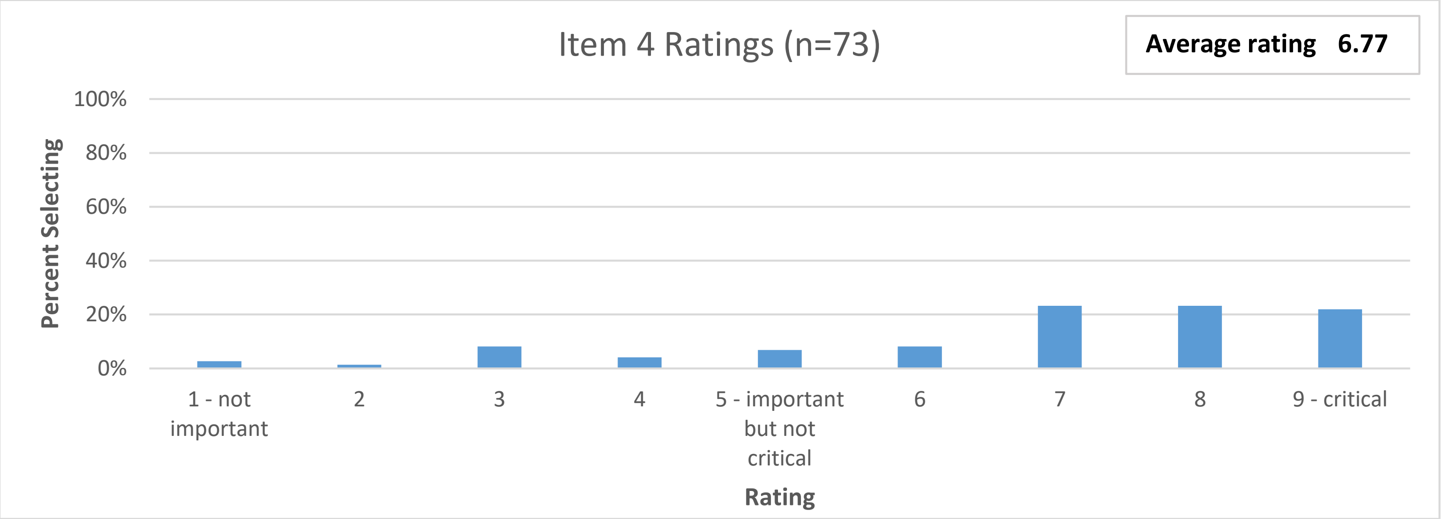

**Comments in favour of importance**

- "This always needs to be defined to interpret the results."
- "Very important, especially for trials with clusters starting the intervention at different time points (such as, but not limited to, stepped wedge trials)"
- "Exposure time can vary so greatly it seems essential to describe."
- "This may not be applicable for all CRTs, but is likely applicable for many (or most), and it would be useful to force investigators to think through and explain this aspect."

**Comments against importance**

- "This will not be an issue for all cluster RCTs, either because exposure time is constant or the effects are instant."
- "This is not solely a feature for CRTs; the same could happen in individually randomised trials with learning/time dependent effects."
- "This is only relevant to some CRTs"

*Item 5*

**Whether treatment effects are marginal or cluster-specific**

*Explanation:* In CRTs, investigators can estimate either marginal or cluster-specific treatment effects. Marginal treatment effects are obtained by summarising potential outcomes for each treatment condition across the whole population and contrasting these summaries. Cluster-specific effects are obtained by contrasting summaries within each cluster first, and then taking an average of the cluster-specific effects.

### Results

73 participants responded to this question; 3 selected ‘unable to rate’.

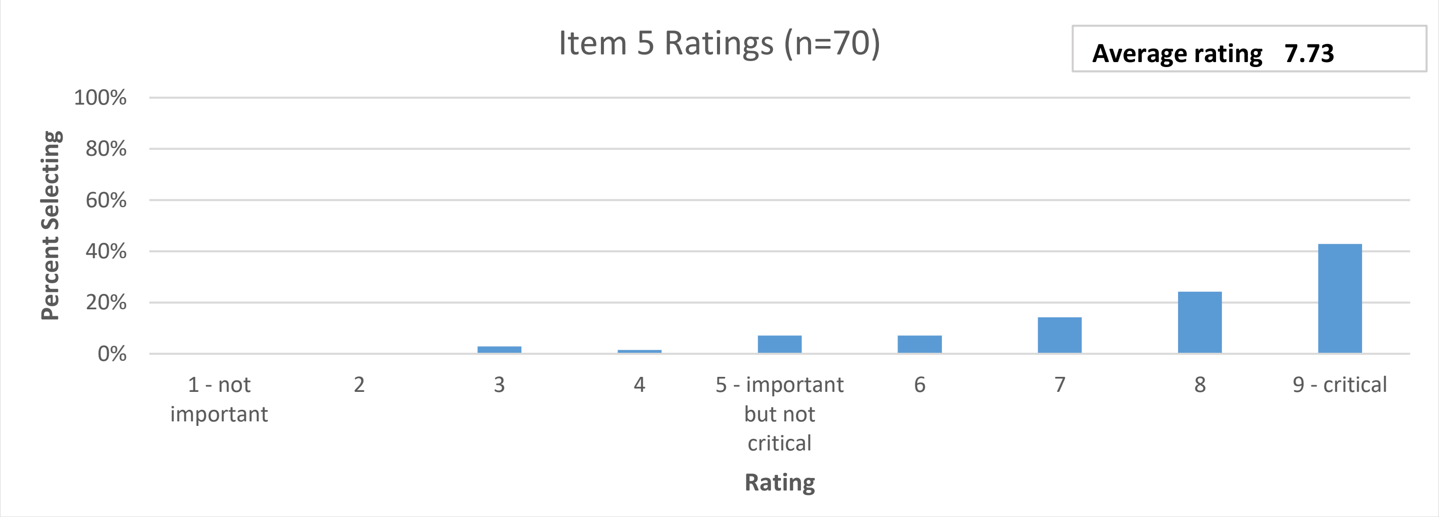

**Comments in favour of importance**

- “Choosing and communicating [this] is essential, especially when different choices can result in different estimates and different interpretations.”
- “It's important to specify whether treatment effects are marginal- or cluster-specific, because they answer different research questions.”
- “This has historically been a big issue in CRTs, so it would be very odd to have guidance on estimands for CRTs that does not include this aspect.”
- “This is critical part of cluster RCTs that's often misunderstood and under appreciated.”

**Comments against importance**

- Only relevant for non-collapsible measures such as odds ratios; for differences, the interpretation of both approaches is the same.
- “The consequences of this choice only makes a difference in some trials; the differences are usually small enough that it’s a second order concern versus more consequential decisions.”
- “I'm not sure about this item. If there were empirical evidence that these estimates often diverge, then I would rate this item more importantly.”

*Item 6*

**Strategies for handling cluster-level intercurrent events**

**Explanation:** Intercurrent events are events occurring after treatment assignment that affect either the interpretation or the existence of the measurements associated with the clinical question of interest (such as changing from the control to intervention treatment or stopping assigned treatment early).

In individually randomised trials, most intercurrent events happen at the individual-level, e.g., an individual stops taking their assigned treatment. However, in CRTs, intercurrent events can also occur at the cluster-level; for instance, a cluster may decide not to implement their assigned treatment, or may delay its implementation for several months. Cluster-level intercurrent events may affect all individuals enrolled in the cluster for the duration of the event.

Just like for individual-level intercurrent events, investigators can estimate different types of treatment effects depending on which strategies they use for cluster-level events (e.g., treatment policy, hypothetical).

### Results

73 participants responded to this question; 1 selected 'Unable to rate'.

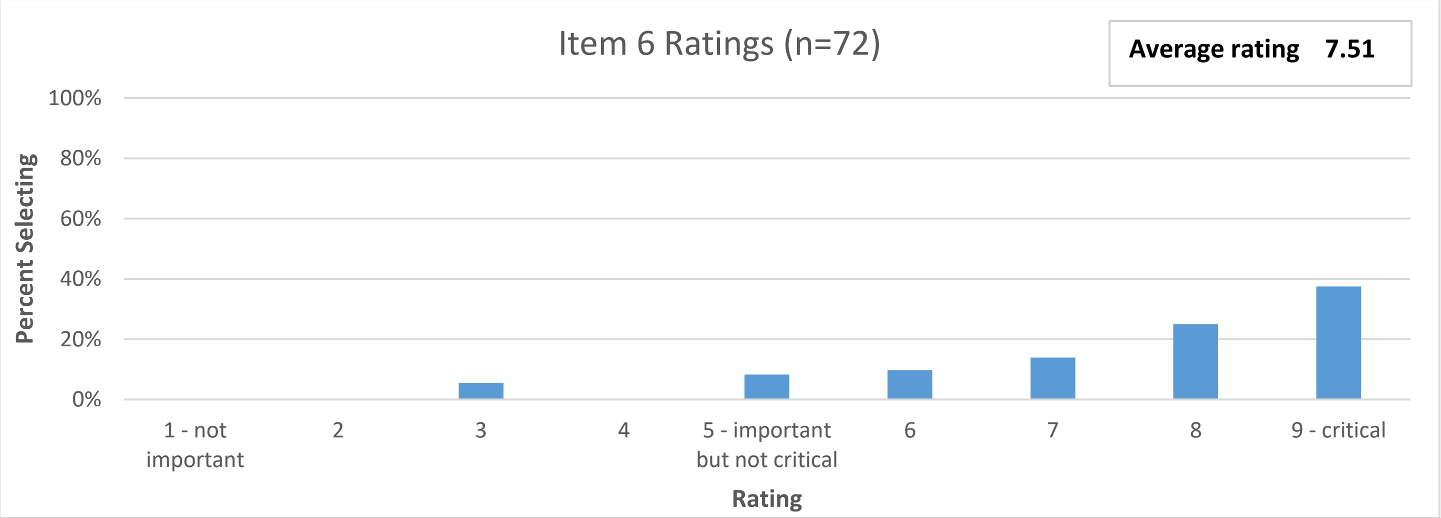

**Comments in favour of importance**

- "What strategy is used to handle intercurrent events … needs to be articulated"
- "This is really important - non-adherence at the cluster level is common (e.g. for many complex interventions, hospitals/clusters implement the intervention later than they should, or don't implement it at all). If this isn't an explicit item in the guidance there's a major risk that investigators don't explain it."
- "Absolutely critical to include all relevant intercurrent events and to distinguish between cluster and individual-level."
- "If one wishes to really understand the effect estimate, one cannot ignore the frequency and types of intercurrent events."

**Comments against importance**

- "It is really hard to anticipate such eventualities and thus preplan strategies."
- "I don't think this is very common in practice. And since such events may be difficult to anticipate in advance (and may be of many different kinds) it may be difficult to pre-define a meaningful estimand."

*Item 7*

**How interference or spillover effects (where treatment of one participant may impact the outcomes of other participants) is handled**

*Explanation:* In some CRTs the treatment one individual receives may affect the outcomes of other individuals. For instance, in a CRT evaluating a vaccine to protect against influenza, if one individual receives the vaccine, this may reduce the chances that other individuals (including those who refused the treatment or do not comply) become infected by reducing the overall level of influenza circulating.

Interference or spillover can occur both within clusters (where individuals who refuse treatment may still receive some benefit due to the individuals who do receive treatment) or between clusters (e.g., if vaccination in a treated cluster reduces influenza prevalence such that individuals in nearby control-arm clusters have less risk of becoming infected).

When interference or spillover occurs, trial investigators could estimate different types of treatment effects, such as the indirect effect of treatment in those who do not receive treatment themselves, or the total effect of treatment which measures both the individual effect of receiving treatment as well as the effect of other individuals in the population receiving the treatment.

### Results

71 participants responded to this question; 2 selected ‘unable to rate’; 2 did not respond.

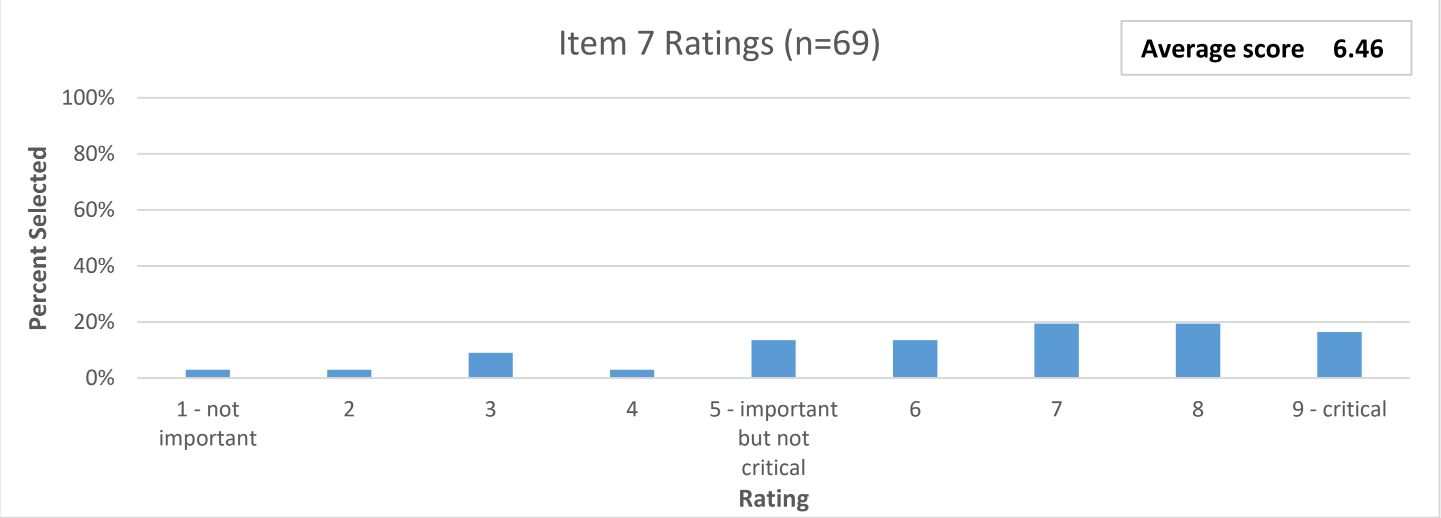

**Comments in favour of importance**

- “We often want to estimate things at the cluster level and so interaction between individuals is to be expected. For education trials one can also get synergy, whereby education works better when given to a group.”
- “Yes, this is critical and present for the study of most infectious diseases and also behavioral/information interventions that permeate a network via discussion processes or network norms. CRTs allow for possible spillover within the study clusters and this is implicitly captured when we index the potential outcomes by the cluster-level exposure. I view it as a missed opportunity in CRTs [not] to evaluate spillover; however, the study would need to capture individual exposure/adherence as well. This also raises a question if CRTs should be powered for spillover if that is an important feature of the intervention.”
- “This is important, but only for certain types of trial or intervention.”

**Comments against importance**

- “This issue seems fairly specific to vaccine trials (I can't think of another setting where this would arise) so I don't think it's that important to include in general guidance.”
- “This affects only a small minority of CRTs, and is thus not very applicable. Furthermore, it's a fairly complex issue that most investigators won't be familiar with and will find difficult to specify; as such, there's a danger that including it in the guidance will make the guidance less relevant due to difficulty of use.”
- “I don’t think this guidance should focus on complicated estimands (e.g. multilevel mediation) when there is so much confusion on defining simple estimands (e.g. cluster vs. participant-average effects).”

*Item 8*

**How individuals who leave or change clusters should be handled**

*Explanation:* In some CRTs, individuals may leave or change clusters during the course of a study. For example, in a CRT taking place in schools, some students may change to a different school or drop out of school entirely. Then, individuals may have belonged to multiple clusters during the study, or belong to no cluster at study's end.

This has implications for the definition of the population of individuals of interest (for instance, whether interest lies in any individuals who belonged to relevant clusters at any point, or only individuals belonging to relevant clusters at a specific endline timepoint), as well as how cluster-specific treatment effects (see Item 5) are defined.

### Results

71 participants responded to this question; 1 selected 'unable to score'; 2 did not respond

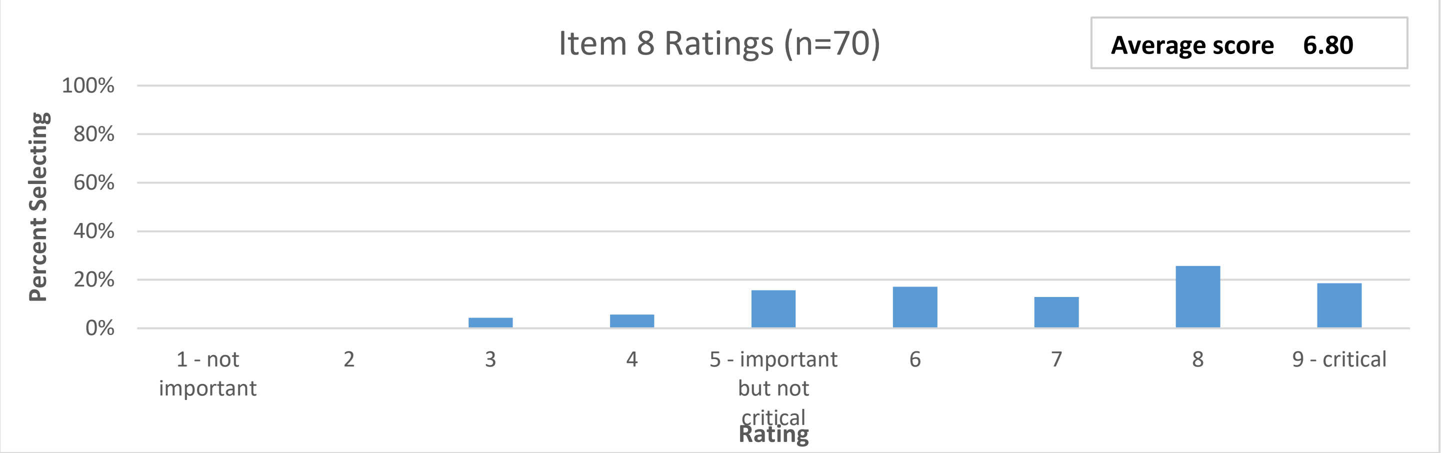

**Comments in favour of importance**

- "This becomes more important depending on design, such as transitioning in or out of schools."
- "I think that it is a critical issue of estimand for CRT, particularly when the cluster have "open population" such as schools or nursing homes."
- "This item is unlikely to apply to all (or possibly most) CRTs. However, for the trial it does apply to it will be quite important."
- "I scored this a little lower just because it's commonly not relevant to the specific trial scenario. However, leaving/changing clusters is an important individual-level intercurrent event and so this needs to be included."

**Comments against importance**

- "I guess this depends on the anticipated level of change. Most cluster trials I have been involved with have had minimal changeover rates."
- "Not widely applicable."
- "I imagine this would come under intercurrent events. A treatment policy strategy would analyse the data according to the intended clusters, for example. ICH e9 does talk about switching treatments and drop out, though unclear how to handle those who switch clusters in the same treatment, so this may be an additional consideration."
- "I'm not sure how big of an issue this is in practice. The problem seems interesting statistically but I'm unclear on how common this occurs, and the magnitude of any bias that may result from individuals leaving/changing clusters."

*New Item: Item 9*

This is a new item developed in response to suggestions by participants during Round 1. You will be asked to rate it as part of Round 2.

**Item 9: How clusters that split, merge, or are empty should be handled**

*Explanation:* In some CRTs, some clusters may split apart into multiple separate clusters during the course of the study. Conversely, some distinct clusters might merge together, or some clusters may remain empty (i.e. include no patients from the target population).

For example, in a CRT where hospitals act as clusters, two hospitals in the same city may merge together, such that the patient sets in both are combined. Another hospital might split into two distinct units, where patients are likewise split across both. Finally, some hospitals may not see any eligible patients from the target population during the study, and so are 'empty'.

These issues may invoke challenges around what constitutes a cluster, and which clusters patients belong to. They may also have implications for how other possible aspects of the estimand are defined, for instance how individuals or clusters are weighted (item 1); the population of clusters (item 2); or how cluster-specific treatment effects are defined (item 5).

*CRT Estimands Extension Delphi: Round 2 Executive Summary*

This executive summary presents the results from Round 2 of the CRT Estimands Extension Delphi, with data collected from November 20-December 11, 2024.

**Sixty-two individuals completed the Round 2 survey (response rate of 85%).**

The summary includes:

In Round 2 of the Delphi, participants again rated the eight items that were rated in Round 1. They also rated and optionally provided comments on a new item, Item 9. In Round 3, you will be asked to rate Item 9 a second (and final) time. You will not re-rate any other items. Therefore, **we present the Round 2 results for Item 9 first**, followed by other items and comments.

*Item 9 (new item for Round 2; to be rated in Round 3)*

**How clusters that split, merge, or are empty should be handled**

*Explanation:* In some CRTs, some clusters may split apart into multiple separate clusters during the course of the study. Conversely, some distinct clusters might merge together, or some clusters may remain empty (i.e. include no patients from the target population).

For example, in a CRT where hospitals act as clusters, two hospitals in the same city may merge together, such that the patient sets in both are combined. Another hospital might split into two distinct units, where patients are likewise split across both. Finally, some hospitals may not see any eligible patients from the target population during the study, and so are 'empty'.

These issues may invoke challenges around what constitutes a cluster, and which clusters patients belong to. They may also have implications for how other possible aspects of the estimand are defined, for instance how individuals or clusters are weighted (item 1); the population of clusters (item 2); or how cluster-specific treatment effects are defined (item 5).

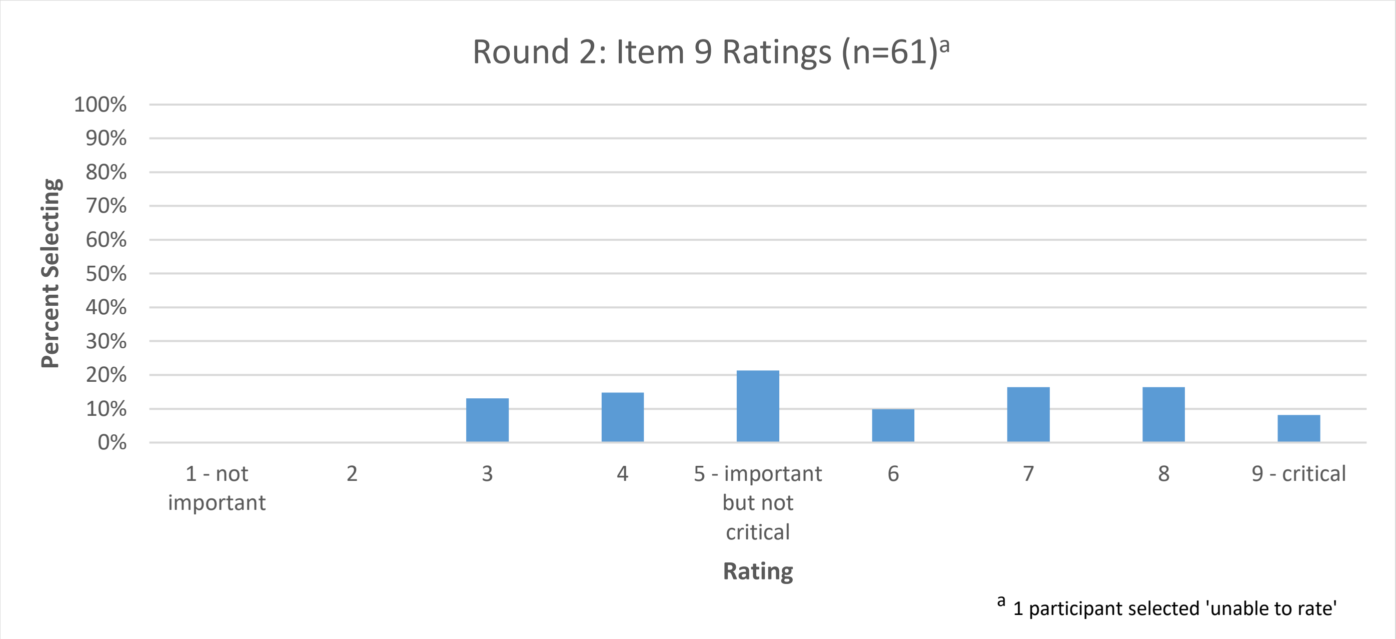

The summary for Item 9 includes a selection of participant comments from Round 2. For those who are interested, a document containing all participant comments for this item can be accessed here.

**Comments in favour of importance**

- "This is very important in terms of understanding exactly how the intervention effect has been calculated, especially when there are multiple options for dealing with this scenario. The situation may not occur very often but it's important to acknowledge this when it does."
- "This seems to be very important as this would potentially change the definition of some clusters and affect the interpretations"

**Comments against importance**

- "This does not seem common enough to merit inclusion in the guidance."
- "Seems like this is a very idiosyncratic that would apply in only a small number of settings."
- "This would presumably be an intercurrent event at the cluster level, I'm not sure it merits a separate item."
- "I think this information is important to convey in a statistical analysis plan (e.g., what defines a cluster and how are these events to be handled in the analysis), but I'm not sure about incorporation into an estimand defined at trial commencement (where such events are unlikely to be planned & may be difficult to foresee)."

*Item 1*

**How individuals and clusters are weighted in the estimand (i.e., individual- or cluster-average treatment effect).**

*Explanation:* In CRTs, individuals and clusters can be weighted differently. For instance, equal weight could be given either to each individual (individual-average effect) or to each cluster (cluster-average effect). These different treatment effects have different interpretations and may lead to different results.

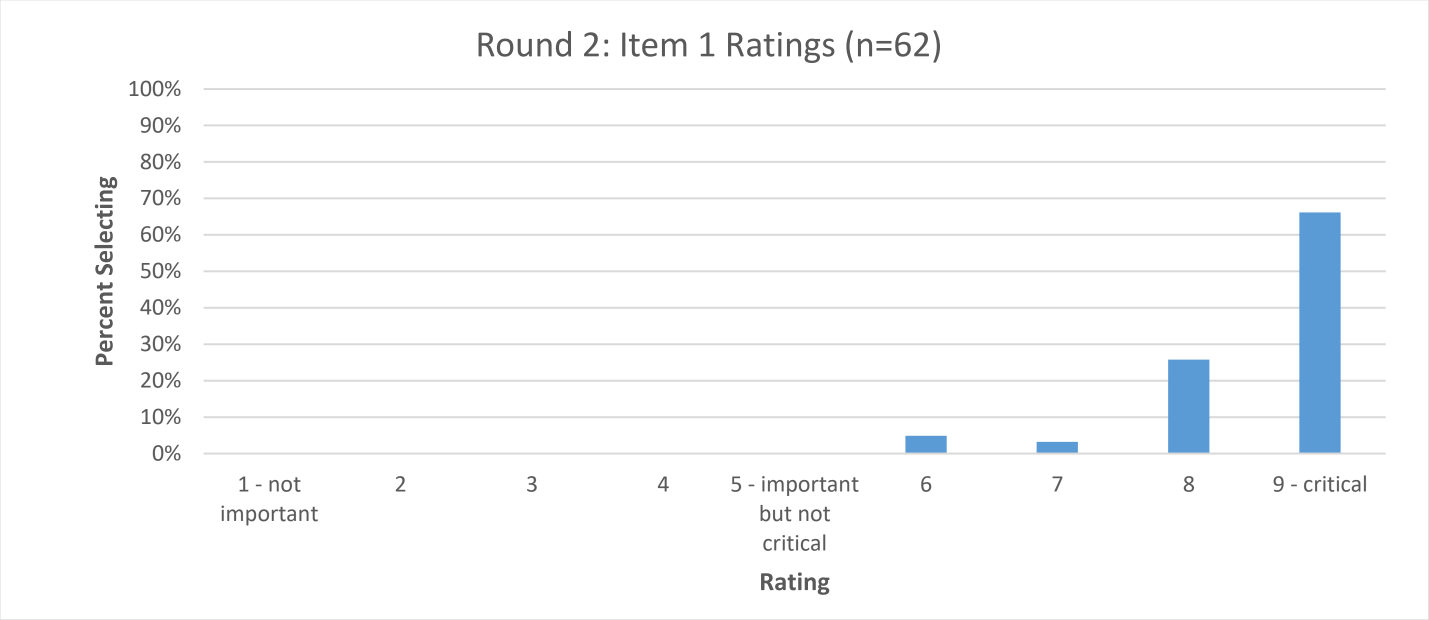

*Item 2*

**Population of clusters**

*Explanation:* The benefit and safety of an intervention may depend on the population for which it is intended. The population of individuals of interest is one of the key estimand attributes listed in the ICH E9(R1) addendum. However, in CRTs, investigators could also choose to estimate the treatment effect for different populations of clusters, such as:

- Specialist hospitals with high-expertise clinicians and resources
- General hospitals, irrespective of the type of clinicians or resources
- The subset of hospitals that would implement the intervention correctly if assigned to the intervention arm

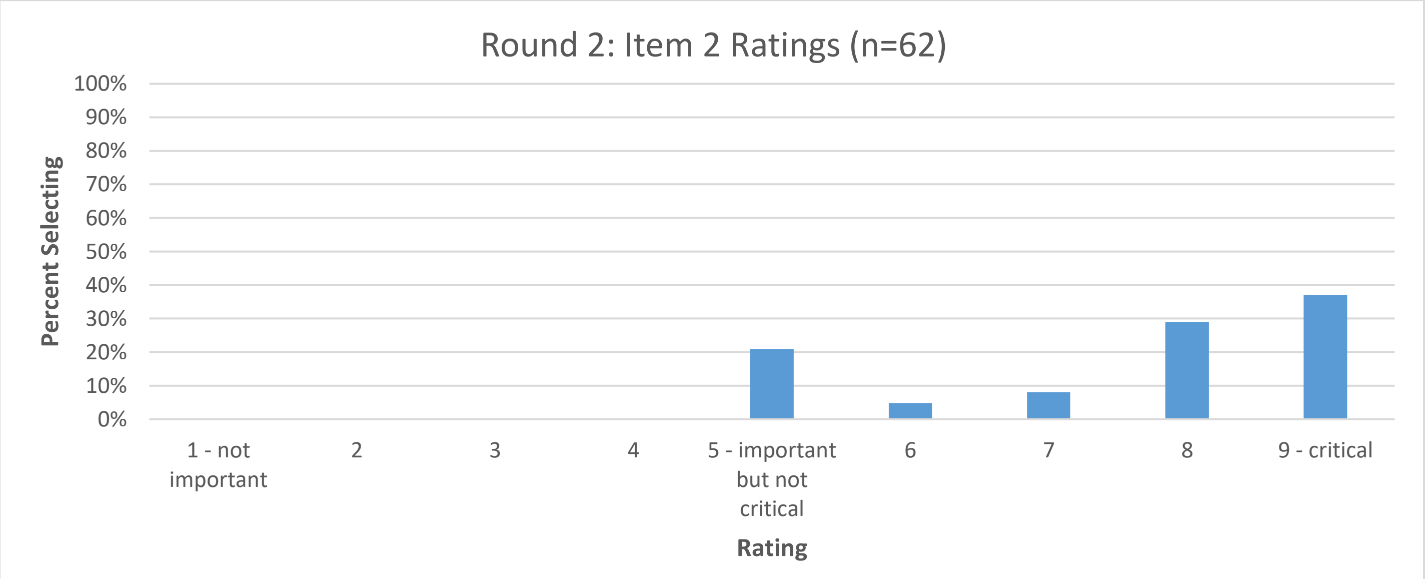

*Item 3*

**Population of individuals under selection or recruitment bias**

*Explanation:* In some CRTs, individuals are enrolled in the trial after randomisation of the clusters. Because individuals or recruiters may be aware of the cluster's treatment assignment, there can lead to selection or recruitment bias, where some individuals may enrol into the trial if their cluster were assigned the intervention but not if it were assigned the control.

In this situation, trial investigators could choose to estimate the treatment effect for different populations of individuals. For instance, they could estimate the effect for all eligible individuals, or for a certain subset defined by whether individuals would enrol according to the cluster treatment assignment or not (e.g., those who would enrol only if their cluster were assigned to the intervention, or those who would enrol regardless of their cluster's treatment assignment).

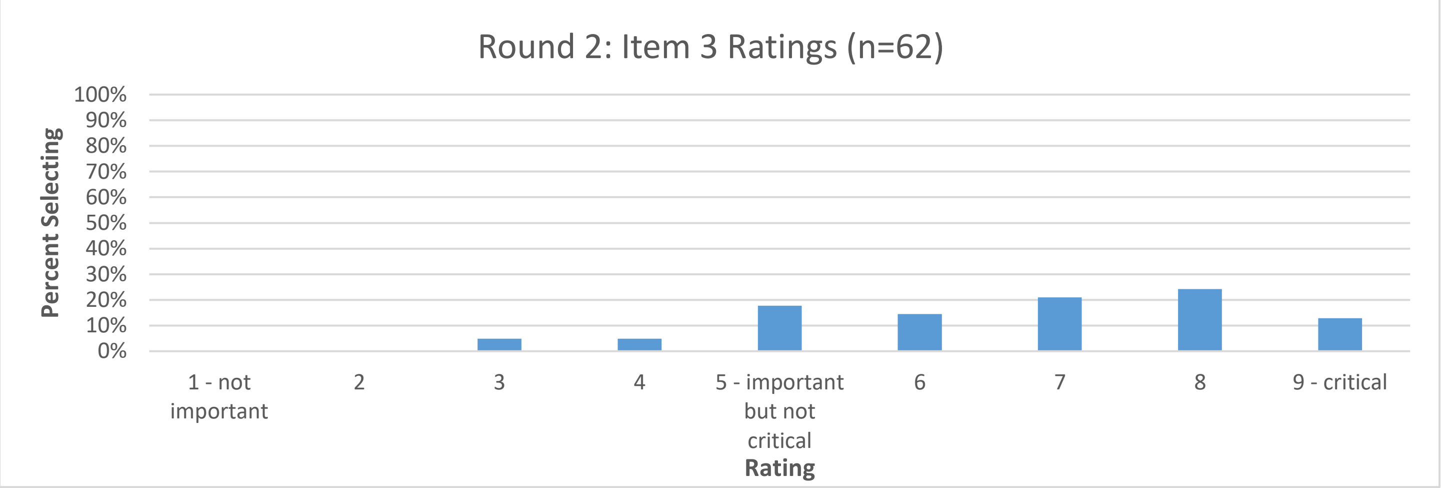

*Item 4*

**Exposure time of clusters and individuals to the intervention**

*Explanation:* In CRTs, both individuals and clusters may be exposed to the intervention for different durations, and the intervention's effect may depend on this exposure time. For example:

- An audit-and-feedback intervention to change clinicians' prescribing behaviour may require time to fully change clinical behaviour, so individuals who enrol a short time after the intervention was first implemented in a cluster may experience less benefit than individuals who enrol a long time after it was implemented.

When the effect of an intervention depends on the exposure time of individuals and/or clusters, trial investigators can estimate different types of treatment effects, such as the treatment effect averaged across different levels of exposure time, or the effect for a specific exposure time (for both individual and cluster-level exposure times).

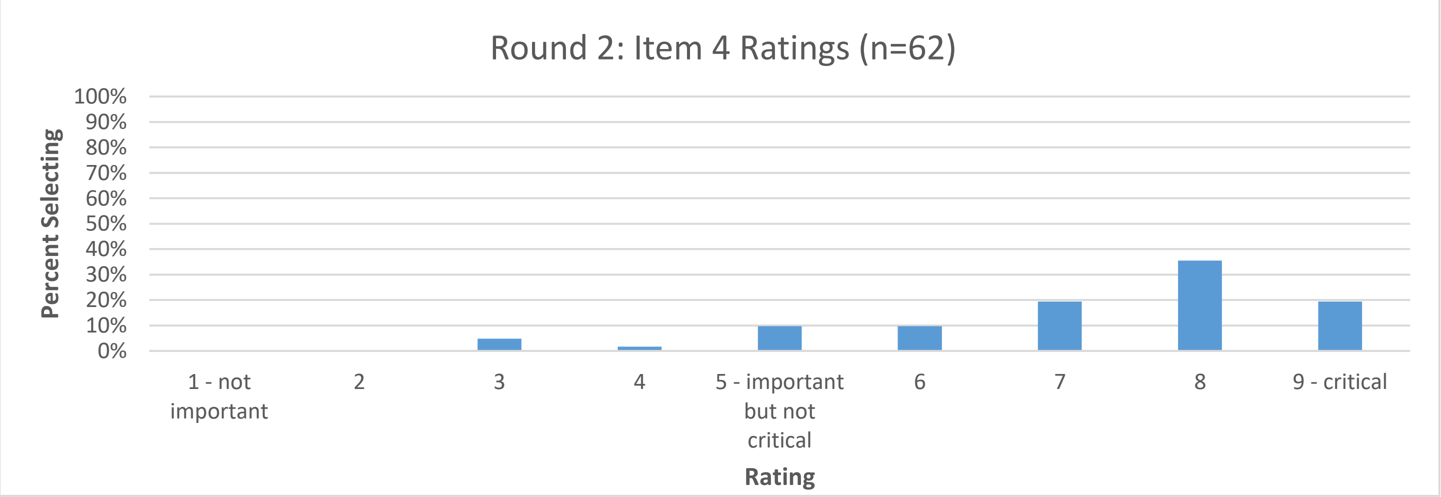

*Item 5*

**Whether treatment effects are marginal or cluster-specific**

*Explanation:* In CRTs, investigators can estimate either marginal or cluster-specific treatment effects. Marginal treatment effects are obtained by summarising potential outcomes for each treatment condition across the whole population and contrasting these summaries. Cluster-specific effects are obtained by contrasting summaries within each cluster first, and then taking an average of the cluster-specific effects.

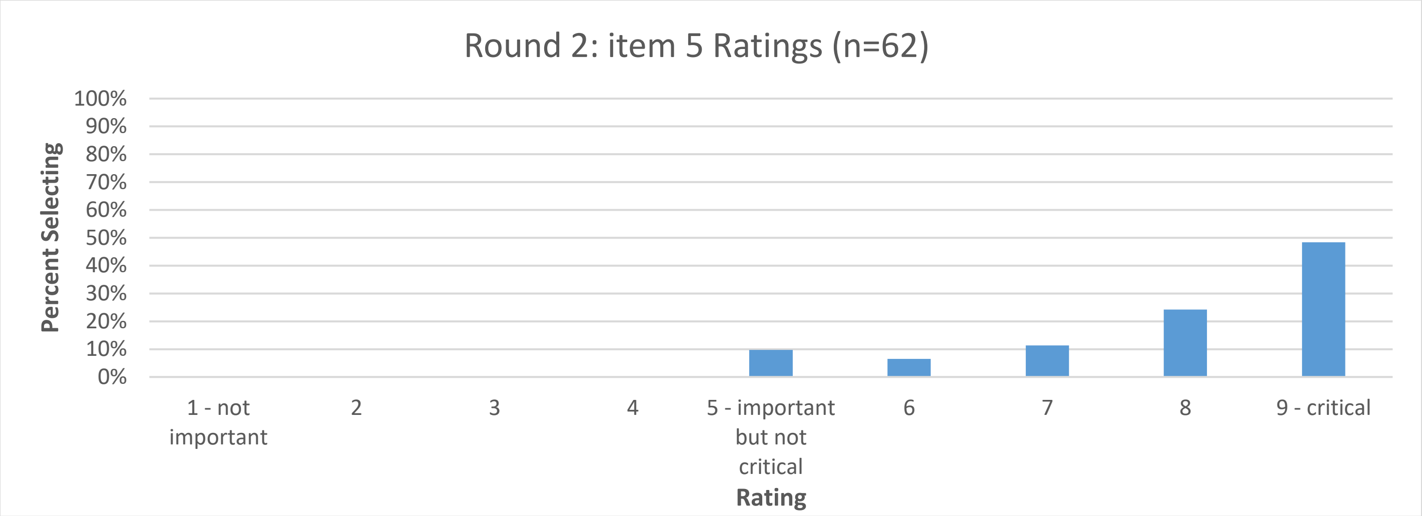

*Item 6*

**Strategies for handling cluster-level intercurrent events**

**Explanation:** Intercurrent events are events occurring after treatment assignment that affect either the interpretation or the existence of the measurements associated with the clinical question of interest (such as changing from the control to intervention treatment or stopping assigned treatment early).

In individually randomised trials, most intercurrent events happen at the individual-level, e.g., an individual stops taking their assigned treatment. However, in CRTs, intercurrent events can also occur at the cluster-level; for instance, a cluster may decide not to implement their assigned treatment, or may delay its implementation for several months. Cluster-level intercurrent events may affect all individuals enrolled in the cluster for the duration of the event.

Just like for individual-level intercurrent events, investigators can estimate different types of treatment effects depending on which strategies they use for cluster-level events (e.g., treatment policy, hypothetical).

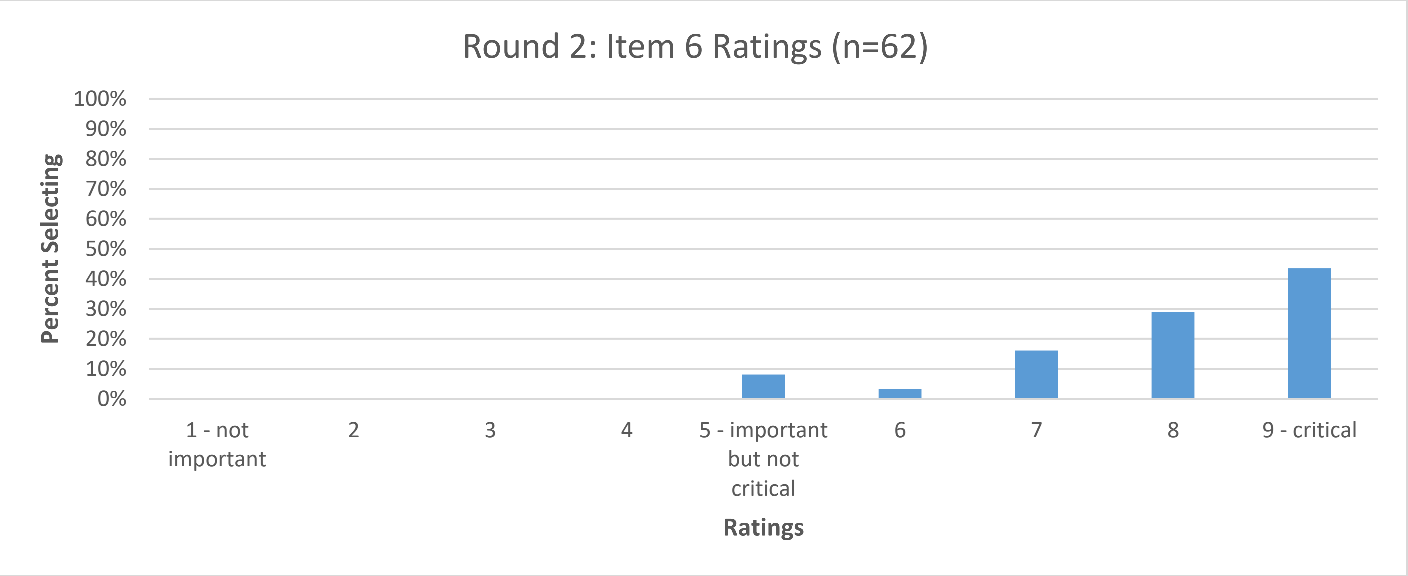

*Item 7*

**How interference or spillover effects (where treatment of one participant may impact the outcomes of other participants) is handled**

*Explanation:* In some CRTs the treatment one individual receives may affect the outcomes of other individuals. For instance, in a CRT evaluating a vaccine to protect against influenza, if one individual receives the vaccine, this may reduce the chances that other individuals (including those who refused the treatment or do not comply) become infected by reducing the overall level of influenza circulating.

Interference or spillover can occur both within clusters (where individuals who refuse treatment may still receive some benefit due to the individuals who do receive treatment) or between clusters (e.g., if vaccination in a treated cluster reduces influenza prevalence such that individuals in nearby control-arm clusters have less risk of becoming infected).

When interference or spillover occurs, trial investigators could estimate different types of treatment effects, such as the indirect effect of treatment in those who do not receive treatment themselves, or the total effect of treatment which measures both the individual effect of receiving treatment as well as the effect of other individuals in the population receiving the treatment.

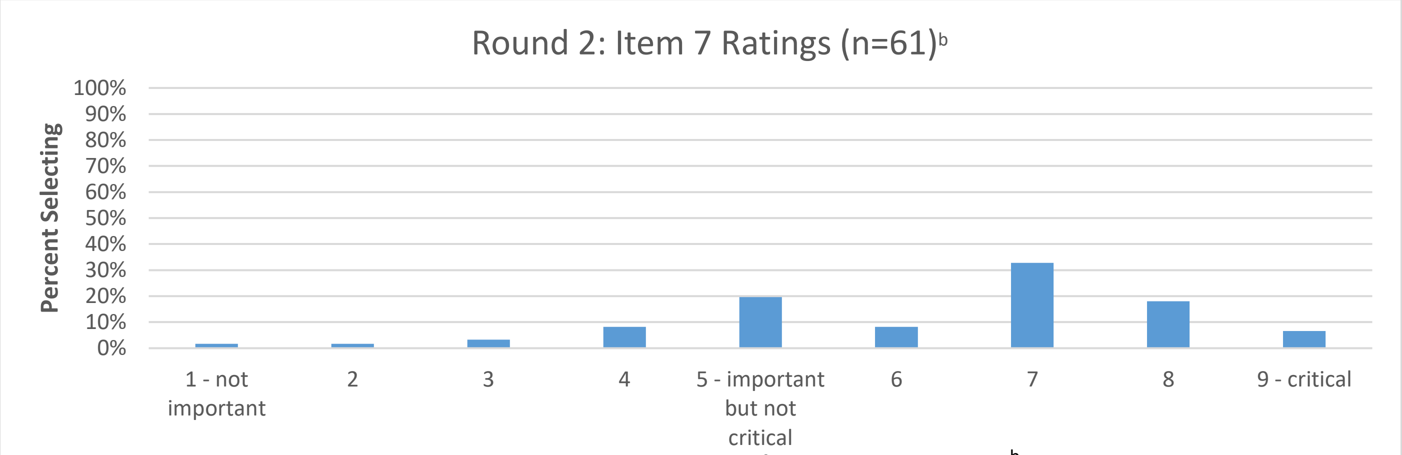

[b] 1 participant selected 'unable to rate'

*Item 8*

**How individuals who leave or change clusters should be handled**

*Explanation:* In some CRTs, individuals may leave or change clusters during the course of a study. For example, in a CRT taking place in schools, some students may change to a different school or drop out of school entirely. Then, individuals may have belonged to multiple clusters during the study, or belong to no cluster at study's end.

This has implications for the definition of the population of individuals of interest (for instance, whether interest lies in any individuals who belonged to relevant clusters at any point, or only individuals belonging to relevant clusters at a specific endline timepoint), as well as how cluster-specific treatment effects (see Item 5) are defined.

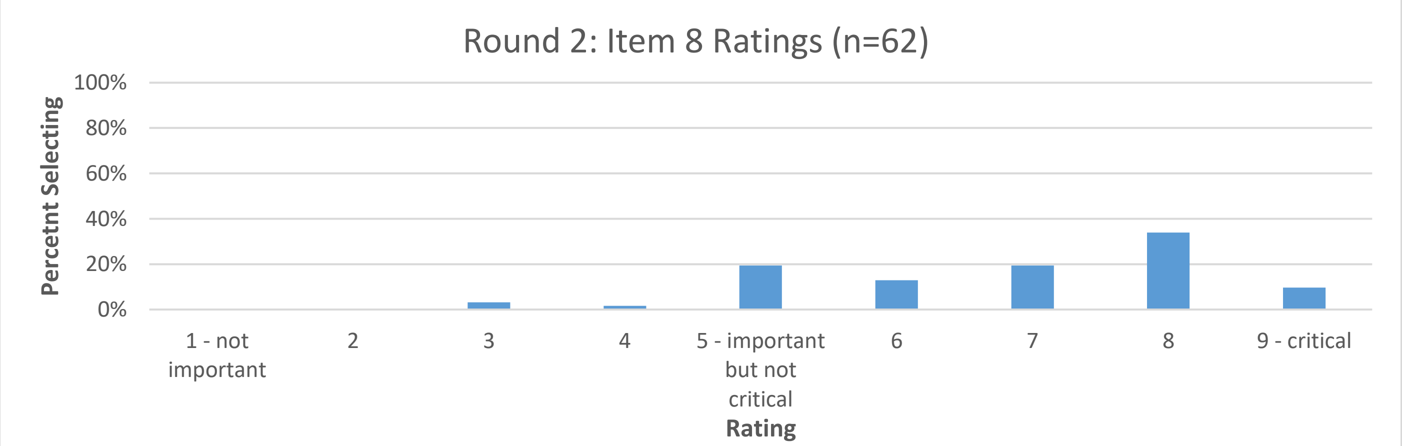

**Table S1. Selected comments from executive summaries**

| | |
|---|---|
| **Item 1: Weighting of individuals and clusters in the estimand (i.e., individual- or cluster-average treatment effect).** | |
| **In favour of importance** | • "Absolutely critical. If clusters are of equal size, this does not matter, but this latter situation is an exception in CRTs not the rule."<br>• "This is fundamentally important to understanding what is being estimated, except in rare situations where the sample size is the same in all clusters."<br>• "It is critical because you can get quite different results depending on how individuals and clusters are weighted."<br>• "Critical to define as methods grow more complex; the estimand is less often implied by the analysis method than it used to be."<br>• "This is absolutely essential … because common analysis methods target different estimands it's essential to know which investigators want to be targeting in order to understand whether the analysis is appropriate, and what assumptions it is making." |
| **Against importance** | • Weights in analysis model more important; specifying estimand weights may indicate common analysis models not appropriate<br>• Only relevant if informative cluster size is present (and will not matter if cluster sizes are all equal)<br>• It does not seem to be something clinicians care about or are able to specify in every trial |
| **Item 2: Population of clusters** | |
| **In favour of importance** | • "Both description of individuals of interest and of clusters of interest are key estimand attributes for a CRT"<br>• "This is important to interpreting results"<br>• "This is important because it affects generalisability" |
| **Against importance** | • "It seems to me that this information would already be captured in explaining the population per the existing ICH E9(R1) guidance"<br>• "Any trial report should describe the cluster population." |
| **Item 3: Population of individuals under selection or recruitment bias** | |
| **In favour of importance** | • "Important because it affects the interpretability of results"<br>• "This is central to understanding the ultimate effect estimate reported in the study. Without this context, making sense of results and thinking about using results in new settings is constrained."<br>• "The risk for such selection or recruitment bias can be quite high and that this point should be considered very seriously in CRTs." |
| **Against importance** | • "An important issue for the conduct of some CRTs (not all) but not necessarily an issue to do with estimands. It seems that the "clinical question of interest" would almost always relate to "all eligible individuals" since in future clinical implementation the issue of post-randomisation enrolment would not apply."<br>• "While this is important for trials affected by selection/recruitment bias, it is not relevant for all trials. It is also (in principle) covered by the "Patient population" attribute. Finally, it can be a technically challenging concept, and investigators may find it challenging to implement routinely (which then makes the guidance less useful overall, if people find it difficult to use)." |
| **Item 4: Exposure time of clusters and individuals to the intervention** | |
| **In favour of importance** | • "This always needs to be defined to interpret the results."<br>• "Very important, especially for trials with clusters starting the intervention at different time points (such as, but not limited to, stepped wedge trials)"<br>• "Exposure time can vary so greatly it seems essential to describe."<br>• "This may not be applicable for all CRTs, but is likely applicable for many (or most), and it would be useful to force investigators to think through and explain this aspect." |
| **Against importance** | • "This will not be an issue for all cluster RCTs, either because exposure time is constant or the effects are instant."<br>• "This is not solely a feature for CRTs; the same could happen in individually randomised trials with learning/time dependent effects."<br>• "This is only relevant to some CRTs" |
| **Item 5: Whether treatment effects are marginal or cluster-specific** | |
| **In favour of importance** | • "Choosing and communicating [this] is essential, especially when different choices can result in different estimates and different interpretations."<br>• "It's important to specify whether treatment effects are marginal- or cluster-specific, because they answer different research questions."<br>• "This has historically been a big issue in CRTs, so it would be very odd to have guidance on estimands for CRTs that does not include this aspect."<br>• "This is critical part of cluster RCTs that's often misunderstood and under appreciated." |

| | |
|---|---|
| **Against importance** | • Only relevant for non-collapsible measures such as odds ratios; for differences, the interpretation of both approaches is the same.<br>• "The consequences of this choice only makes a difference in some trials; the differences are usually small enough that it's a second order concern versus more consequential decisions."<br>• "I'm not sure about this item. If there were empirical evidence that these estimates often diverge, then I would rate this item more importantly." |
| **Item 6: Handling of cluster-level intercurrent events** | |
| **In favour of importance** | • "What strategy is used to handle intercurrent events … needs to be articulated"<br>• "This is really important - non-adherence at the cluster level is common (e.g. for many complex interventions, hospitals/clusters implement the intervention later than they should, or don't implement it at all). If this isn't an explicit item in the guidance there's a major risk that investigators don't explain it."<br>• "Absolutely critical to include all relevant intercurrent events and to distinguish between cluster and individual-level."<br>• "If one wishes to really understand the effect estimate, one cannot ignore the frequency and types of intercurrent events." |
| **Against importance** | • "It is really hard to anticipate such eventualities and thus preplan strategies."<br>• "I don't think this is very common in practice. And since such events may be difficult to anticipate in advance (and may be of many different kinds) it may be difficult to pre-define a meaningful estimand." |
| **Item 7: Handling of interference or spillover effects** | |
| **In favour of importance** | • "We often want to estimate things at the cluster level and so interaction between individuals is to be expected. For education trials one can also get synergy, whereby education works better when given to a group."<br>• "Yes, this is critical and present for the study of most infectious diseases and also behavioral/information interventions that permeate a network via discussion processes or network norms. CRTs allow for possible spillover within the study clusters and this is implicitly captured when we index the potential outcomes by the cluster-level exposure. I view it as a missed opportunity in CRTs [not] to evaluate spillover; however, the study would need to capture individual exposure/adherence as well. This also raises a question if CRTs should be powered for spillover if that is an important feature of the intervention."<br>• "This is important, but only for certain types of trial or intervention." |
| **Against importance** | • "This issue seems fairly specific to vaccine trials (I can't think of another setting where this would arise) so I don't think it's that important to include in general guidance."<br>• "This affects only a small minority of CRTs, and is thus not very applicable. Furthermore, it's a fairly complex issue that most investigators won't be familiar with and will find difficult to specify; as such, there's a danger that including it in the guidance will make the guidance less relevant due to difficulty of use."<br>• "I don't think this guidance should focus on complicated estimands (e.g. multilevel mediation) when there is so much confusion on defining simple estimands (e.g. cluster vs. participant-average effects)." |
| **Item 8: Handling of individuals who leave or change clusters** | |
| **In favour of importance** | • "This becomes more important depending on design, such as transitioning in or out of schools."<br>• "I think that it is a critical issue of estimand for CRT, particularly when the cluster have "open population" such as schools or nursing homes."<br>• "This item is unlikely to apply to all (or possibly most) CRTs. However, for the trial it does apply to it will be quite important."<br>• "I scored this a little lower just because it's commonly not relevant to the specific trial scenario. However, leaving/changing clusters is an important individual-level intercurrent event and so this needs to be included." |
| **Against importance** | • "I guess this depends on the anticipated level of change. Most cluster trials I have been involved with have had minimal changeover rates."<br>• "Not widely applicable."<br>• "I imagine this would come under intercurrent events. A treatment policy strategy would analyse the data according to the intended clusters, for example. ICH e9 does talk about switching treatments and drop out, though unclear how to handle those who switch clusters in the same treatment, so this may be an additional consideration."<br>• "I'm not sure how big of an issue this is in practice. The problem seems interesting statistically but I'm unclear on how common this occurs, and the magnitude of any bias that may result from individuals leaving/changing clusters." |
| **Item 9: Handling of clusters that split, merge, or are empty** | |
| **In favour of importance** | • "This is very important in terms of understanding exactly how the intervention effect has been calculated, especially when there are multiple options for dealing with this scenario. The situation may not occur very often but it's important to acknowledge this when it does." |

| | |
|---|---|
| | • "This seems to be very important as this would potentially change the definition of some clusters and affect the interpretations" |
| **Against importance** | • "This does not seem common enough to merit inclusion in the guidance."<br>• "Seems like this is a very idiosyncratic that would apply in only a small number of settings."<br>• "This would presumably be an intercurrent event at the cluster level, I'm not sure it merits a separate item."<br>• "I think this information is important to convey in a statistical analysis plan (e.g., what defines a cluster and how are these events to be handled in the analysis), but I'm not sure about incorporation into an estimand defined at trial commencement (where such events are unlikely to be planned & may be difficult to foresee)." |
